# Exploring possible point-of-sale restrictions for vapes and nicotine pouches: cross-sectional surveys with adults and adolescents in the UK

**DOI:** 10.64898/2026.09.23.26363788

**Authors:** Anne Marie MacKintosh, Catherine Best, Crawford Moodie, Kathryn Angus, Georgia Alexandrou, Allison Ford

## Abstract

**Introduction:** Within the Tobacco and Vapes Act 2026, the UK Government is considering restricting retail displays of vaping and other nicotine products to discourage youth uptake. We explored how adult nicotine users and adolescents responded to conventional and restricted displays.

**Methods:** Two UK-wide cross-sectional online surveys were conducted April-June 2026, with current and former adult nicotine users (n=2,851), and adolescents (11 17 years; n=2,123). Participants rated images of vape and nicotine pouch point-of-sale (POS) displays (two conventional, two restricted) across 10 items. Other measures included demographics, smoking/vaping/nicotine pouch use, vaping susceptibility, and product choice.

**Results:** Ratings of two conventional POS images were consistently favourable. Two restricted POS images were almost exclusively rated unfavourably. GEE analyses, controlling for demographics and behaviour, indicated that a display with standardised white packs (versus a conventional display) consistently had lower odds of favourable ratings (adults range:0.13-0.31; adolescents range:0.11-0.32). A fully covered display had even lower odds of favourable ratings on all items (adults range:0.06-0.19; adolescents range:0.05-0.10). Presented with hypothetical restricted displays, most adults using nicotine remained consistent with their product choice. Favourable ratings of conventional displays were positively associated with adolescents’ vaping susceptibility. Those favourably rating a large, branded display were more likely to be susceptible than those giving unfavourable/neutral ratings (AOR=2.73, 95%CI [2.16-3.45] p<0.001). No association was found between favourable ratings of restricted displays and susceptibility.

**Conclusion:** Restricting POS displays could reduce positive messaging around vaping and adolescent susceptibility. The potential impact of restrictions on adult users’ perceptions of harm and accessibility should be considered.

**What is already known on this topic:**

- Open point-of-sale (POS) displays are an effective marketing tool which increase attention, communicate positive product attributes, influence social norms, and drive sales.
- While evidence demonstrates the impact of open and banned POS display of tobacco, there is a lack of research on how adults and adolescents respond to conventional and restricted POS displays of vapes or other nicotine products,

**What this study adds:**

- Adults and adolescents almost exclusively rated conventional POS images favourably, and restricted POS images, unfavourably.
- Favourable ratings of conventional displays were positively associated with adolescents’ vaping susceptibility, while no association was found between favourable ratings of restricted displays and susceptibility.

**How this study might affect research, practice or policy:**

- Restricting POS displays may reduce positive vaping messages and adolescents’ susceptibility to vaping,

## INTRODUCTION

As part of the United Kingdom (UK) Tobacco and Vapes Act 2026,^1^ the UK Government is considering restricting the retail display of vaping and other nicotine products.^2^ Citing concerns over youth vaping and nicotine pouch use,^3,4^ and the potential for nicotine addiction and harms,^5^ restricting the open display of vaping and nicotine products is aimed at discouraging young people from using these products through reducing their visibility and appeal.^6^

Point-of-sale (POS) displays are an established marketing strategy for retailers to effectively communicate with consumers. A display with a massed set of packs is described as having a ‘billboarding effect’ and being a significant form of advertising for companies.^7^ Display size, composition, presentation and product assortment send signals to consumers on product attributes such as popularity, quality, value and price.^8^ Research across consumer goods demonstrates that displays capture and increase attention and interest in products, influence purchase decision-making processes, encourage trial, and drive sales.^8,9^ Larger, and more organised, attractive, and relevant displays are particularly important for encouraging impulse purchasing.^7,8,10,11^. Displays also influence normative evaluations. By communicating norms around others’ purchases and behaviour, displays can indicate socially desirable activities and reassure consumers on their own purchases.^10–12^

Banning open POS displays, as many jurisdictions have done for tobacco,^13^ reduces the ability for companies to communicate their product at POS. Evaluations of tobacco display bans have consistently found that bans reduce POS marketing exposure in smokers and non-smokers,^14–17^ and help denormalise smoking.^14,18^ For people who smoke, display bans result in fewer impulse buys^15,19^ and support quitting.^20,21^ For young people, bans reduce smoking initiation and susceptibility to smoke.^16–18,21,22^

There is limited research on actual or perceived consumer response to restrictions on the display of vapes or other nicotine products, possibly because many countries promptly banned it.^2^ Studies with adolescents in the UK^23^ and Scotland^24^ found that exposure to a greater number of vape POS display images increased susceptibility to smoking among those who visited retail stores more regularly,^23^ and an association between recall of vape POS displays and use of, and intention to use, vapes.^24^ A longitudinal study with adolescents in the US found that recall of vape POS displays predicted susceptibility to vape across a range of store types.^25^ A UK-wide survey found that young people were more likely than adults to notice vapes displayed in a shop window and to have positive views about them.^26^ In a qualitative study, both adults and adolescents felt that brightly coloured and large window displays signalled that vaping was targeted towards children and young people and moved their perception of vapes away from a potential smoking cessation aid.^26^

Retail trade magazines highlight recent investment in vape and nicotine pouch displays given they are viewed as *‘key to driving purchase of next-gen products’* in the UK.^27^ Retailers report dedicating greater space to nicotine products^28^ and being encouraged to sign up for new backwall gantries, such as those offered by British American Tobacco (BAT),^29^ which feature LED lighting around products, digital screens and greater space to increase nicotine pouch visibility. One retailer participating in a 4-week trial of BAT’s new gantry reported a 35% increase in sales of BAT’s nicotine pouch brand Velo.^30^ Adolescents aged 11-17 years in Great Britain increasingly report awareness of vape promotion in shops, with it suggested that shop displays may be a driver of increased awareness and uptake of nicotine pouches among adolescents.^3^

To inform the UK Government’s consultation on prohibiting the display of vaping and other nicotine products,^2^ this paper explores how current and former adult nicotine users and adolescents respond to conventional displays and potential display restrictions.

## METHODS

### Design and participants

Two online cross-sectional surveys were conducted with current and former adult nicotine users (18+) and adolescents (11-17 years) in the UK. YouGov, an online survey provider, hosted the surveys using their large existing UK panel. YouGov recruited two non-probability samples, with the adolescent sample seeking to be representative of adolescents in the UK. For adults, YouGov targeted panel members who fulfilled smoking/vaping eligibility criteria. For 11-15-year-olds, panel members known to have a child within this age range were contacted. For 16 17-year-olds, existing panel members were targeted directly, or via parents on the panel. The adult survey was conducted April-May 2026, the adolescent survey April-June 2026. Ethical approval was granted by the University of Stirling General University Ethics Panel (GUEP 2025 23794 18341).

### Procedure

For adults and adolescents (16+), a survey link was emailed to eligible panel members with the information sheet and consent form. For adolescents (11 15) both parent and participant viewed their own information and consent forms. Informed consent was necessary to proceed to the survey. YouGov undertook quality checks to ensure integrity of the final data. Cases suspected of giving false answers, cases with inconsistencies in recency of nicotine use as well as cases missing demographic information were excluded.

### Development and testing

Survey items were informed by research on tobacco POS and packaging,^16,31,32^ and six public involvement discussion groups, four with adults who smoked, vaped and/or used nicotine pouches, and two with adolescents (14-15, 16-17). Within groups participants discussed their reactions to images of vape and nicotine pouch displays, to identify suitable images for the survey, and draft measures for assessing response to displays. Drafts of questions and response options were tested and refined in 16 individual cognitive interviews with adults and adolescents (11-17 years) to ensure comprehension, relevance and acceptability.

### Point-of-sale display images

There were two conventional display images: Image (a) (large display of vapes and nicotine pouches without tobacco) and Image (b) (display of vapes and nicotine pouches beside covered tobacco), and two restricted displays images: Image (c) (display of vapes and nicotine pouches in white standardised packaging^33^ beside covered tobacco) and Image (d) (display of covered vapes and nicotine pouches beside covered tobacco), see Figure 1. Images were based on photographs of a UK in-store display (taken with retailer consent) adapted using Adobe Photoshop and an artificial intelligence (AI) tool (ChatGPT).

**Figure 1:**
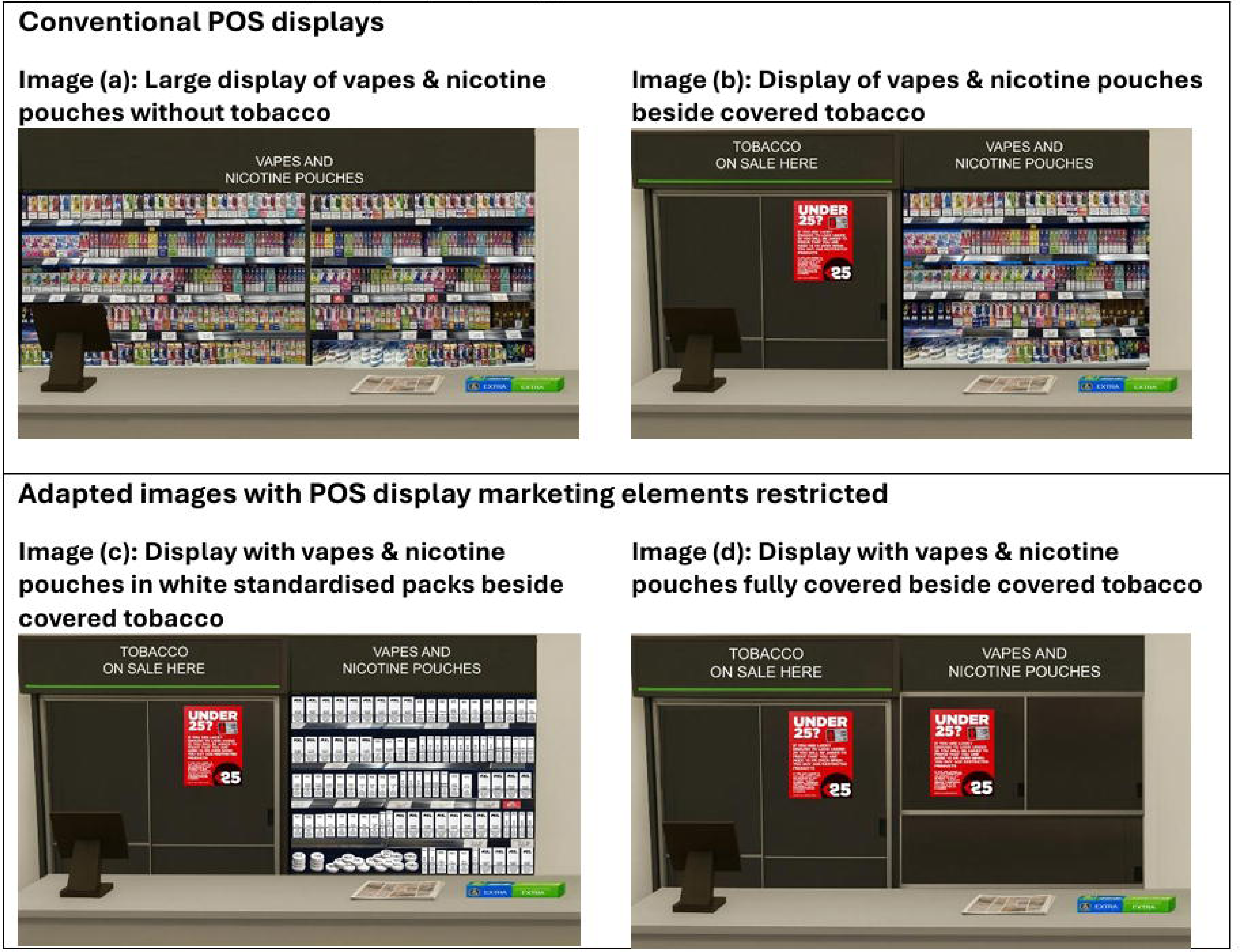
Point-of-sale (POS) display images.

## Measures

### Demographics

Age, gender, social grade and country were obtained from YouGov. Social grade was determined by the occupation of the chief income earner in the household (ABC1=middle class, C2DE=working class).

### Smoking and vaping status

Questions about product use were supported with product images to avoid ambiguity. Smoking status was determined with ‘Which of the following best describes you? (1) I have never smoked, not even tried a puff or two, (2) I tried smoking in the past but I never smoke cigarettes (including hand-rolled) now, (3) I used to smoke but I never smoke cigarettes (including hand-rolled) now, (4) I smoke cigarettes (including hand-rolled) but not every day, (5) I smoke cigarettes (including hand-rolled) every day’. Those responding 4-5 were categorised as currently smoking.

Vaping status was determined with ‘Which of these best describes whether or not you have ever used or tried vapes? (1) I have never used vapes, (2) I have only ever tried vapes once or twice, (3) I have used vapes in the past but I never use them now, (4) I occasionally use vapes (less than once a month), (5) I use vapes at least once a month, (6) I use vapes at least once a week, (7) I use vapes every day’. Those responding 4-7 were categorised as currently vaping.

A four-category classification of vaping/smoking status was established from these questions: (1) Dual use, (2) Exclusively vapes, (3) Exclusively smokes, and (4) Does not currently vape/smoke.

### Nicotine pouch use

Adults were asked about nicotine pouch use with response options from ‘I have never used nicotine pouches’ to ‘I use nicotine pouches ever day’ and ‘Not sure’. Adolescents were asked whether they had ever used pouches (Yes, No, Not sure). Ever users were asked when they last used a nicotine pouch. Those indicating use in the past 30 days were classed as currently using.

### Adolescents’ susceptibility to vape

Susceptibility was assessed across three items, adapted from past smoking and vaping research.^34–37^ Never vapers were classed as non-susceptible if they answered ‘Definitely not’ to the questions: ‘If one of your friends offered you a vape, would you use it?’ and ‘Do you think you will try or use a vape at any time during the next year?’ and to the likelihood that ‘you will be using vapes when 18 years old’.

### Display responses

Ten items assessed responses to the four POS images (Figure 1). Participants were asked: ‘Please give the number that best describes what you think about the way vapes are displayed in this picture’ and were assessed via five-point semantic scales, with a ‘Not sure’ option: (1) Makes vaping seem appealing/Makes vaping seem unappealing; (2) Looks eye-catching/Does not look eye-catching; (3) Makes me think that it’s OK to vape/Makes me think that it’s not OK to vape; (4) Makes it feel like vapes are meant for someone like me/Makes it feel like vapes are not meant for someone like me; (5) Makes it easy for someone like me to buy vapes/Makes it difficult for someone like me to buy vapes; (6) Makes me think that vapes are not at all harmful to health/ Makes me think that vapes are very harmful to health; (7) Tempts me to vape/ Puts me off vaping; (8) Makes me think that lots of people vape/ Makes me think that hardly anyone vapes; (9) Looks attractive/ Looks unattractive; (10) Makes vaping seem fun/ Makes vaping seem boring. A random split sample ensured half received the negative statement first and half the positive statement. Items were coded to make a low score (1) indicative of a negative/unfavourable assessment and a high score (5) indicative of a positive/favourable assessment.

### Product choice

For Images (b) to (d) (Figure 1), adults who currently used nicotine were asked: ‘Imagine you are looking to buy a nicotine product and the shop you go into has this display. Which product, if any, would you be most likely to choose from this display? (Please assume the brand and product you want is available.)’ Response options were: (1) Cigarettes/tobacco; (2) Vapes/vaping products; (3) Nicotine pouches (sometimes called snus-style nicotine pouches); (4) None of these; (5) Not sure.

### Control variables

Adolescents were asked about peer, sibling and parental vaping.

### Statistical analysis

Data analysis was undertaken using SPSSv31. Adult and adolescent samples were analysed separately but the approach to analysis was the same for both, with control variables for multivariable analysis tailored appropriately. Descriptive statistics for the adolescent sample are weighted to reflect the population of 11–17-year-olds in the UK. Multivariable analyses are unweighted as these controlled for key demographic variables. The adult sample is unweighted as it is not intended to be representative.

Paired t-tests were used to generate mean scores for each of the ten items for each POS image. A total score for the ten items was also calculated for each image, with Cronbach’s alpha exceeding 0.88 for each. Image (b) was selected as the reference category for all comparisons as it represented a conventional display where tobacco is sold alongside vapes and pouches. Mean scores for each of the other images were assessed relative to Image (b). Due to the ordinal nature of the data from the five-point scales, the Wilcoxon signed rank test, a non-parametric procedure suited to paired data, was used to test for significant differences between ratings. Effect sizes (r) were estimated from the Wilcoxon signed rank results using the formula r=|Z|/√N.^38^ Bonferroni adjustments were incorporated to account for the multiple comparisons, resulting in a critical alpha of 0.017.

Ratings were also dichotomised, with 4-5 classed as favourable and 1-3 neutral/unfavourable. For the total scores, 31-50 was classed as favourable and 10-30 neutral/unfavourable. As the same participants rated four images, the analysis also had to account for repeated measures and the correlation between individual participants’ ratings of the four images. Generalised estimating equations (GEE) for binary outcomes were used to produce estimates of the likelihood of favourable ratings, comparing the POS images while accounting for repeated measures. An exchangeable correlation structure was used. The model outcome variable was whether a favourable rating was given (e.g. looks eye-catching). Image (b) was again used as the reference category, and the GEE analysis provided a sensitivity analysis for the Wilcoxon signed rank tests on the ten items. The GEE analysis controlled for gender (female vs. male); age group (adults 55+ and 35-54 vs. 18-34; adolescents 17, 15-16 and 13-14 vs. 11-12); social grade (C2DE vs. ABC1); current vaping and/or smoking status (exclusively smoke, exclusively vape and dual use vs. neither vape nor smoke currently) and current use of nicotine pouches (currently use vs. does not use).

Among adolescents who had never vaped, logistic regressions were run, for each image, to assess any potential association between overall favourable rating of each image and susceptibility. These controlled for: age (17, 15 16 and 13 14 vs. 11 12); gender (female vs. male); social grade (C2DE vs. ABC1); peer vaping (any friends vape, not sure vs. no), sibling vaping (any vape, not sure vs. no) and parental vaping (either parent vapes, not sure vs. no).

Product choice was assessed, for each image, using descriptive statistics. Images (c) and (d) were also cross tabulated with (b) to assess consistency or change in selection relative to the reference Image (b).

## RESULTS

A total of 2,851 adults and 2,123 adolescents were included, with respondent characteristics presented in Table 1.

**Table 1:**
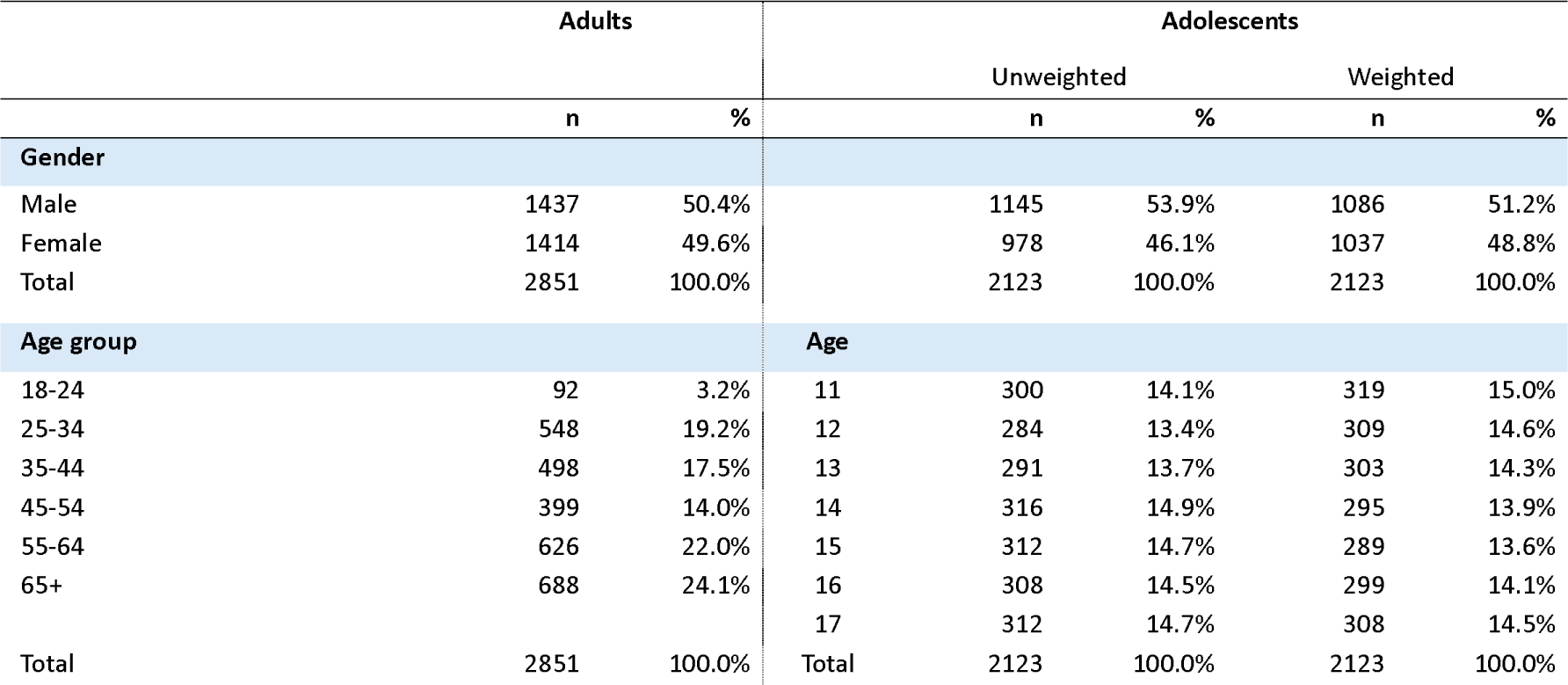
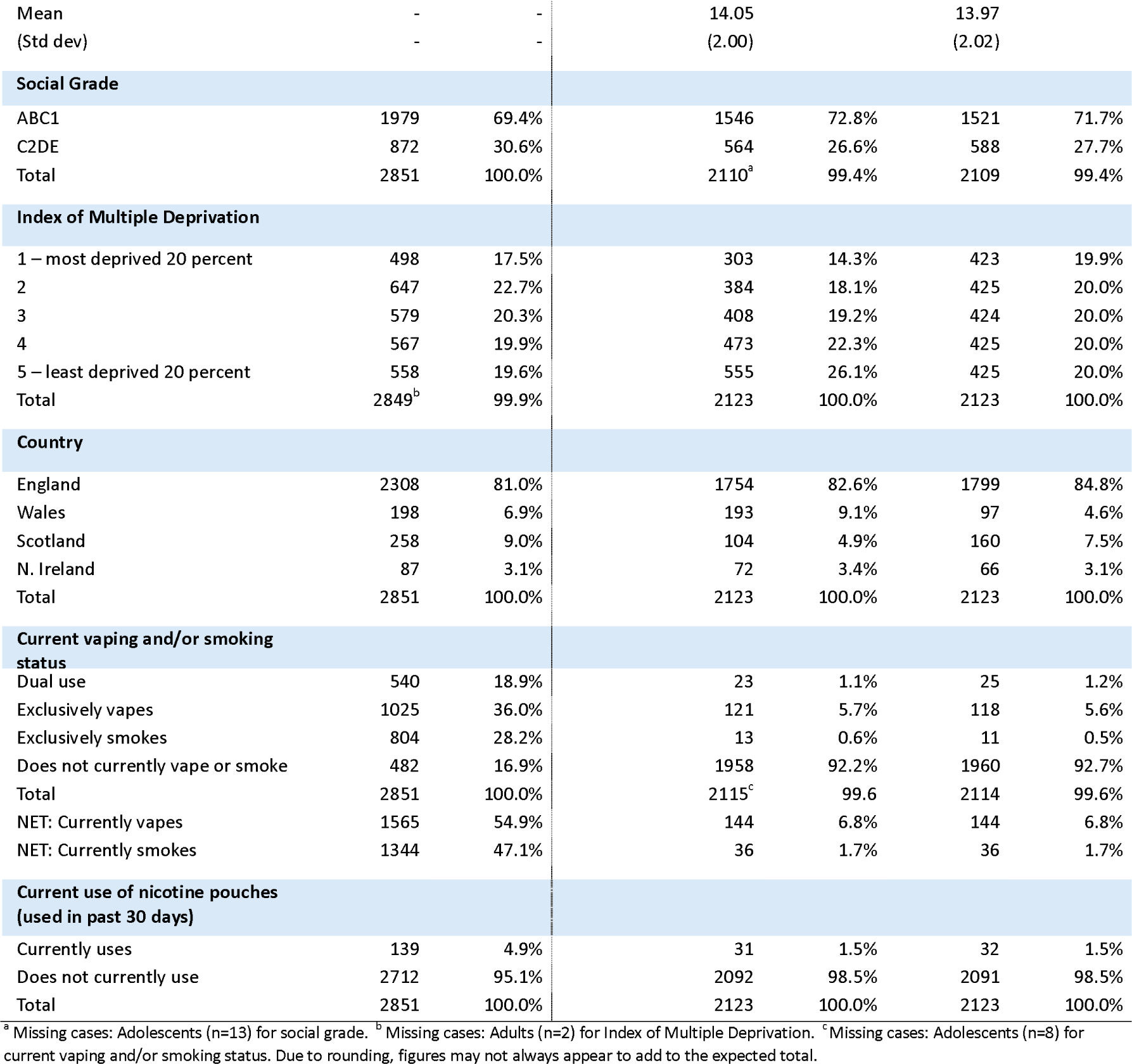
Sample characteristics: Adults (Current and former nicotine users) and Adolescents aged 11 to 17 years.

### POS Image (a): Large display of fully-branded vapes and pouches without tobacco

Adults gave favourable ratings (Figure 2a; Table S1a), with mean scores above the midpoint (3) for all items (range: 3.30-4.21). Most considered this display eye-catching (62.4%), attractive (57.4%) and suggesting that lots of people vape (73.9%) (Table 2a). Most felt it positioned vapes as not being harmful to health (52.9%), vaping as being fun (54.9%) and it being OK to vape (53.6%). In terms of receptivity, around two-fifths felt the display made vaping seem appealing (42.0%), tempted them to vape (42.4%) and suggested vapes are meant for someone like them (38.6%). At a functional level, this display was seen as making it easy to buy vapes (68.8%).

**Figure 2a:**
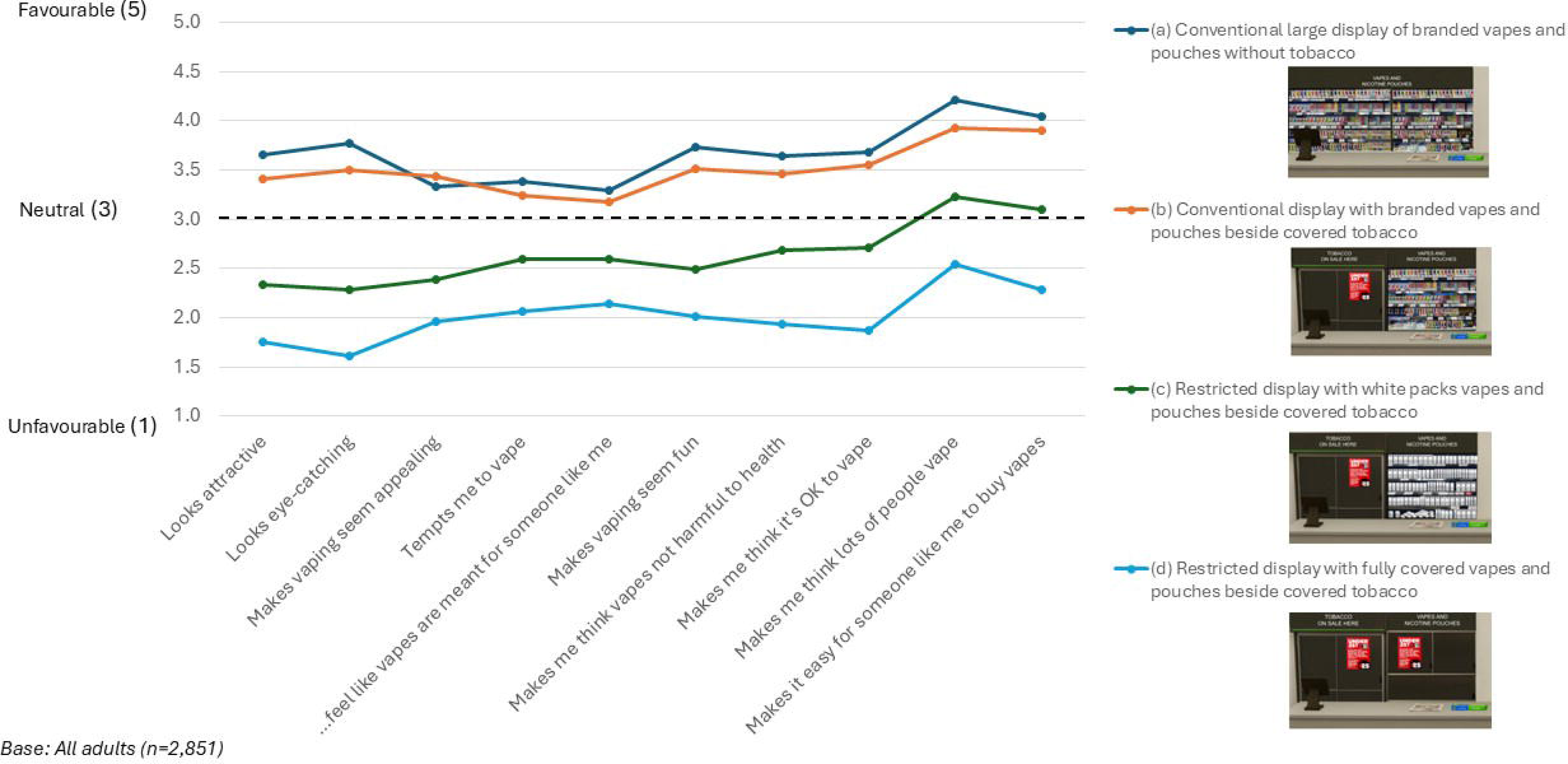
Mean ratings on PoS images (Adults)

**Table 2a.**
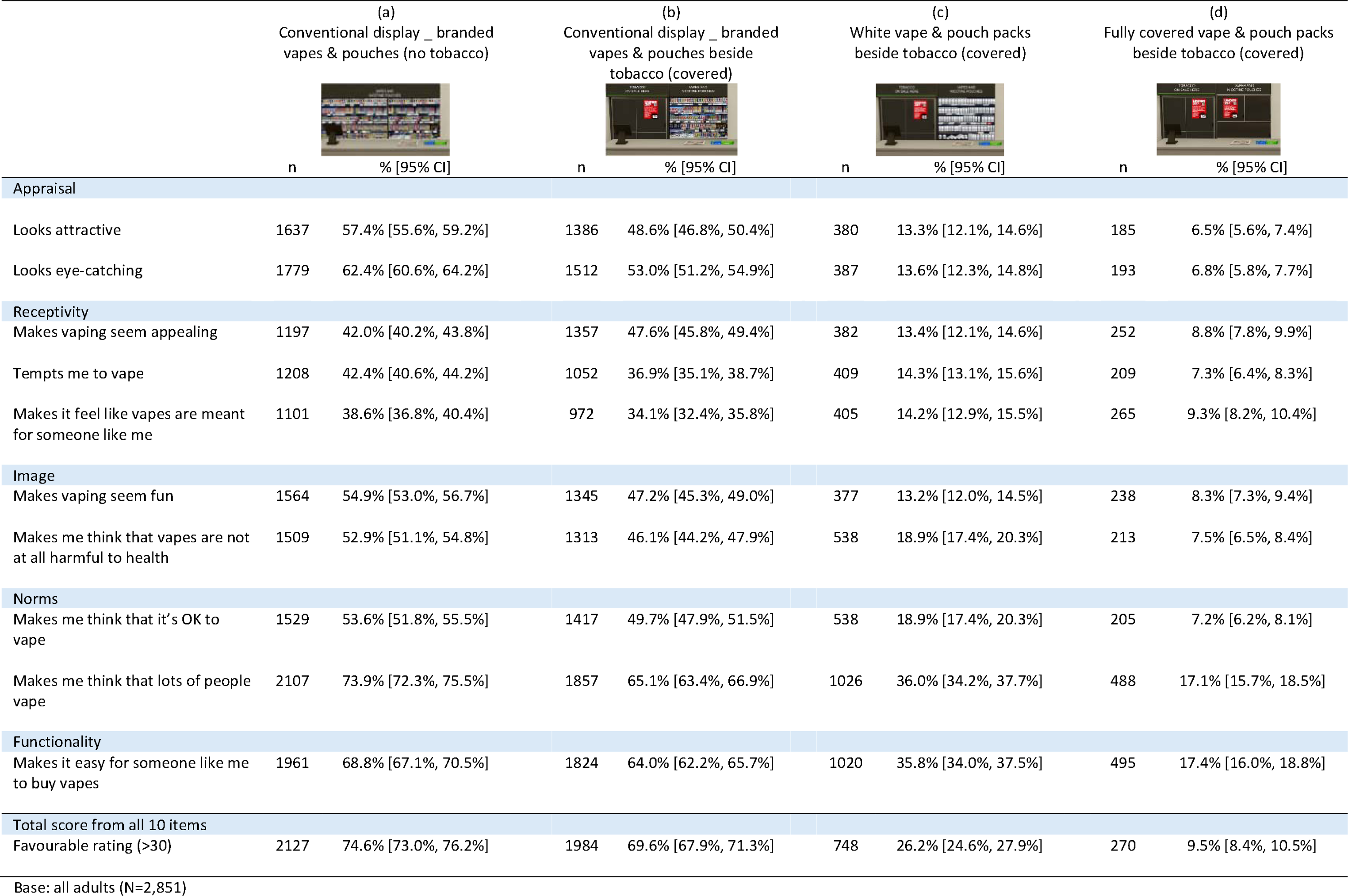
Proportion of adults giving favourable ratings (scores 4 and 5) on POS images.

|  | (a)<br>Conventional display _ branded<br>vapes & pouches (no tobacco) |  | (b)<br>Conventional display _ branded<br>vapes & pouches beside<br>tobacco (covered) |  | (c)<br>White vape & pouch packs<br>beside tobacco (covered) |  | (d)<br>Fully covered vape & pouch packs<br>beside tobacco (covered) |  |
| --- | --- | --- | --- | --- | --- | --- | --- | --- |
|  | n | % [95% CI] | n | % [95% CI] | n | % [95% CI] | n | % [95% CI] |
| **Appraisal** |  |  |  |  |  |  |  |  |
| Looks attractive | 1637 | 57.4% [55.6%, 59.2%] | 1386 | 48.6% [46.8%, 50.4%] | 380 | 13.3% [12.1%, 14.6%] | 185 | 6.5% [5.6%, 7.4%] |
| Looks eye-catching | 1779 | 62.4% [60.6%, 64.2%] | 1512 | 53.0% [51.2%, 54.9%] | 387 | 13.6% [12.3%, 14.8%] | 193 | 6.8% [5.8%, 7.7%] |
| **Receptivity** |  |  |  |  |  |  |  |  |
| Makes vaping seem appealing | 1197 | 42.0% [40.2%, 43.8%] | 1357 | 47.6% [45.8%, 49.4%] | 382 | 13.4% [12.1%, 14.6%] | 252 | 8.8% [7.8%, 9.9%] |
| Tempts me to vape | 1208 | 42.4% [40.6%, 44.2%] | 1052 | 36.9% [35.1%, 38.7%] | 409 | 14.3% [13.1%, 15.6%] | 209 | 7.3% [6.4%, 8.3%] |
| Makes it feel like vapes are meant for someone like me | 1101 | 38.6% [36.8%, 40.4%] | 972 | 34.1% [32.4%, 35.8%] | 405 | 14.2% [12.9%, 15.5%] | 265 | 9.3% [8.2%, 10.4%] |
| **Image** |  |  |  |  |  |  |  |  |
| Makes vaping seem fun | 1564 | 54.9% [53.0%, 56.7%] | 1345 | 47.2% [45.3%, 49.0%] | 377 | 13.2% [12.0%, 14.5%] | 238 | 8.3% [7.3%, 9.4%] |
| Makes me think that vapes are not at all harmful to health | 1509 | 52.9% [51.1%, 54.8%] | 1313 | 46.1% [44.2%, 47.9%] | 538 | 18.9% [17.4%, 20.3%] | 213 | 7.5% [6.5%, 8.4%] |
| **Norms** |  |  |  |  |  |  |  |  |
| Makes me think that it's OK to vape | 1529 | 53.6% [51.8%, 55.5%] | 1417 | 49.7% [47.9%, 51.5%] | 538 | 18.9% [17.4%, 20.3%] | 205 | 7.2% [6.2%, 8.1%] |
| Makes me think that lots of people vape | 2107 | 73.9% [72.3%, 75.5%] | 1857 | 65.1% [63.4%, 66.9%] | 1026 | 36.0% [34.2%, 37.7%] | 488 | 17.1% [15.7%, 18.5%] |
| **Functionality** |  |  |  |  |  |  |  |  |
| Makes it easy for someone like me to buy vapes | 1961 | 68.8% [67.1%, 70.5%] | 1824 | 64.0% [62.2%, 65.7%] | 1020 | 35.8% [34.0%, 37.5%] | 495 | 17.4% [16.0%, 18.8%] |
| **Total score from all 10 items** |  |  |  |  |  |  |  |  |
| Favourable rating (>30) | 2127 | 74.6% [73.0%, 76.2%] | 1984 | 69.6% [67.9%, 71.3%] | 748 | 26.2% [24.6%, 27.9%] | 270 | 9.5% [8.4%, 10.5%] |

Base: all adults (N=2,851)

Adolescents also gave favourable ratings (Figure 2b; Table S1b), with means above the midpoint (range: 3.12-4.16) for all items except ‘makes vaping seem appealing’ (2.89, 95%CI [2.83-2.95]). The other receptivity measures, temptation to vape (3.12, 95%CI [3.06-3.18]) and feeling that vaping is meant for them (3.15, 95%CI [3.09-3.21]), were at the upper end of neutral. Even where average ratings were neutral, a third felt the display made vaping seem appealing (33.4%), and around two-fifths indicated it tempted them to vape (40.2%) and conveyed vapes as being meant for someone like them (42.3%) (Table 2b). Most adolescents (50.8% to 74.4%) gave favourable ratings on the other items, with almost three-quarters indicating that the display made them think that lots of people vape (74.4%).

**Figure 2b:**
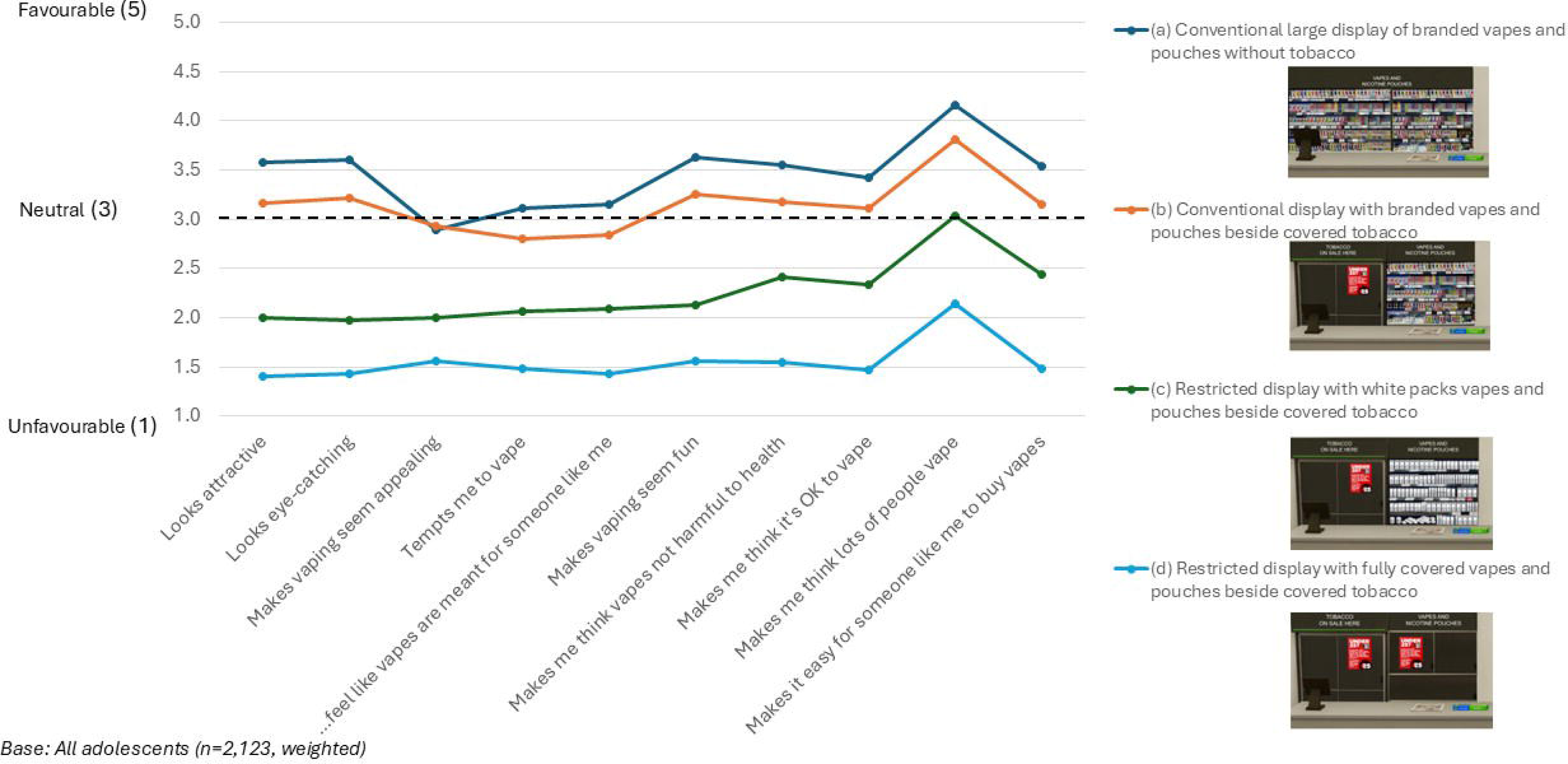
Mean ratings on PoS images (Adolescents)

**Table 2b.**
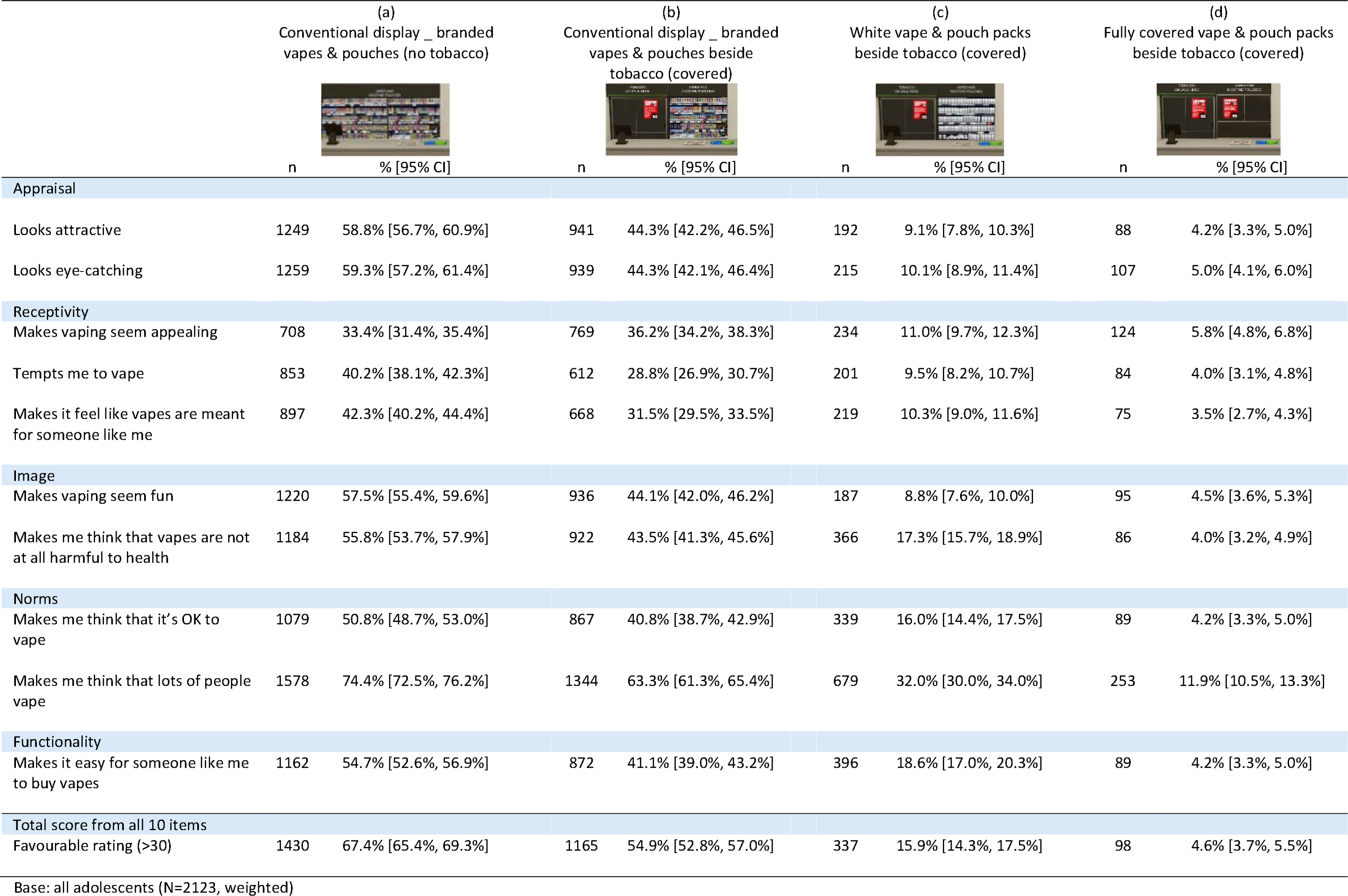
Proportion of adolescents giving favourable ratings (scores 4 and 5) on POS images.

|  | (a)<br>Conventional display _ branded<br>vapes & pouches (no tobacco) |  | (b)<br>Conventional display _ branded<br>vapes & pouches beside<br>tobacco (covered) |  | (c)<br>White vape & pouch packs<br>beside tobacco (covered) |  | (d)<br>Fully covered vape & pouch packs<br>beside tobacco (covered) |  |
| --- | --- | --- | --- | --- | --- | --- | --- | --- |
|  | n | % [95% CI] | n | % [95% CI] | n | % [95% CI] | n | % [95% CI] |
| **Appraisal** |  |  |  |  |  |  |  |  |
| Looks attractive | 1249 | 58.8% [56.7%, 60.9%] | 941 | 44.3% [42.2%, 46.5%] | 192 | 9.1% [7.8%, 10.3%] | 88 | 4.2% [3.3%, 5.0%] |
| Looks eye-catching | 1259 | 59.3% [57.2%, 61.4%] | 939 | 44.3% [42.1%, 46.4%] | 215 | 10.1% [8.9%, 11.4%] | 107 | 5.0% [4.1%, 6.0%] |
| **Receptivity** |  |  |  |  |  |  |  |  |
| Makes vaping seem appealing | 708 | 33.4% [31.4%, 35.4%] | 769 | 36.2% [34.2%, 38.3%] | 234 | 11.0% [9.7%, 12.3%] | 124 | 5.8% [4.8%, 6.8%] |
| Tempts me to vape | 853 | 40.2% [38.1%, 42.3%] | 612 | 28.8% [26.9%, 30.7%] | 201 | 9.5% [8.2%, 10.7%] | 84 | 4.0% [3.1%, 4.8%] |
| Makes it feel like vapes are meant for someone like me | 897 | 42.3% [40.2%, 44.4%] | 668 | 31.5% [29.5%, 33.5%] | 219 | 10.3% [9.0%, 11.6%] | 75 | 3.5% [2.7%, 4.3%] |
| **Image** |  |  |  |  |  |  |  |  |
| Makes vaping seem fun | 1220 | 57.5% [55.4%, 59.6%] | 936 | 44.1% [42.0%, 46.2%] | 187 | 8.8% [7.6%, 10.0%] | 95 | 4.5% [3.6%, 5.3%] |
| Makes me think that vapes are not at all harmful to health | 1184 | 55.8% [53.7%, 57.9%] | 922 | 43.5% [41.3%, 45.6%] | 366 | 17.3% [15.7%, 18.9%] | 86 | 4.0% [3.2%, 4.9%] |
| **Norms** |  |  |  |  |  |  |  |  |
| Makes me think that it's OK to vape | 1079 | 50.8% [48.7%, 53.0%] | 867 | 40.8% [38.7%, 42.9%] | 339 | 16.0% [14.4%, 17.5%] | 89 | 4.2% [3.3%, 5.0%] |
| Makes me think that lots of people vape | 1578 | 74.4% [72.5%, 76.2%] | 1344 | 63.3% [61.3%, 65.4%] | 679 | 32.0% [30.0%, 34.0%] | 253 | 11.9% [10.5%, 13.3%] |
| **Functionality** |  |  |  |  |  |  |  |  |
| Makes it easy for someone like me to buy vapes | 1162 | 54.7% [52.6%, 56.9%] | 872 | 41.1% [39.0%, 43.2%] | 396 | 18.6% [17.0%, 20.3%] | 89 | 4.2% [3.3%, 5.0%] |
| **Total score from all 10 items** |  |  |  |  |  |  |  |  |
| Favourable rating (>30) | 1430 | 67.4% [65.4%, 69.3%] | 1165 | 54.9% [52.8%, 57.0%] | 337 | 15.9% [14.3%, 17.5%] | 98 | 4.6% [3.7%, 5.5%] |

Base: all adolescents (N=2123, weighted)

### POS Image (b): Display with fully-branded vapes and pouches beside covered tobacco

Adults gave favourable ratings, with mean scores above the midpoint (range: 3.18-3.93) for all items (Figure 2a; Table S1a). Most thought it was eye-catching (53.0%), conveyed that lots of people vape (65.1%) and made it easy for buying vapes (64.0%) (Table 2a). Apart from the item on prevalence of vaping (3.81, 95%CI [3.76-3.86]), adolescents gave this image neutral ratings (range: 2.80-3.26) (Figure 2b; Table S1b). They rated six items at the upper end of neutral (range: 3.12-3.26) and the three receptivity items at the lower end of neutral (range: 2.80-2.93). While average ratings were neutral, at least two-fifths considered the display to be attractive (44.3%) and eye-catching (44.3%) and to convey vaping as fun (44.1%) and not harmful to health (43.5%), it being OK to vape (40.8%) and make it easy for someone like them to buy vapes (41.1%) (Table 2b). More than three-fifths thought the image conveyed that lots of people vape (63.3%).

### POS Image (c): Display with vapes and pouches in white standardised packs beside covered tobacco

Adults gave unfavourable ratings for most items, with all but two of the mean scores below the midpoint (range: 2.29-2.71) (Figure 2a; Table S1a). Two items received average neutral ratings: perception that lots of people vape (3.23, 95%CI [3.19-3.27]) and the ease of buying vapes (3.10, 95%CI [3.06-3.15]).

Apart from the perception that lots of people vape (3.04, 95%CI [2.99-3.09]), adolescents gave unfavourable ratings on this display with mean scores all below the midpoint (range: 1.98-2.44) (Figure 2b; Table S1b). Almost a third (32.0%) of adolescents felt this image conveyed that lots of people vape (Table 2b).

### POS Image (d): Display with vapes and pouches fully covered beside covered tobacco

Adults gave unfavourable ratings for all items with mean scores at or below the midpoint (range: 1.61-2.54) (Figure 2a; Table S1a). Fewer than a fifth (17.1%) thought that it suggested lots of people vape while a similar proportion thought it made it easy for buying vapes (17.4%) (Table 2a).

Adolescents also gave unfavourable ratings on this display (range: 1.41-2.14) (Figure 2b; Table S1b) with only 11.9% indicating that this display made them think lots of people vape (Table 2b).

### Comparison of ratings between two conventional POS displays: Image (a) vs. Image (b)

The paired analyses (Table S1a) indicated that, for nine items, adults rated the large fully-branded vape and pouch display (Image a) more favourably than the display of fully-branded vapes and pouches alongside covered tobacco (Image b), see Table S1a. The estimated effect sizes indicated mainly small effects with medium effect sizes (r≥0.3) observed for attractive, eye-catching and perception that lots of people vape.^38^ Adults rated Image (a) less favourably than Image (b) in terms of making vaping seem appealing (p<0.001)

Adolescents also rated Image (a) more favourably than Image (b) on all items (Table S1b) except ‘making vaping seem unappealing/appealing’, which did not show any statistically significant difference within the paired analysis.

The GEE analyses (Table S2a, Table S2b) which took account of repeated measures and controlled for key demographic and behavioural variables verified the above results with Image (a) having higher odds of being rated favourably, for adults (range: 1.17-1.52) and adolescents (range: 1.53-1.88), compared to Image (b), for all items except appeal. Compared with Image (b), adults were less likely to rate Image (a) as appealing (Adjusted Odds Ratio (AOR)=0.79, 95%CI [0.74-0.85] p<0.001).

### Comparison of ratings between a restricted and conventional display: Image (c) vs. Image (b)

Paired analyses indicated that adults (Table S1a) and adolescents (Table S1b) consistently rated the display with white vape and pouch packs (Image c) less favourably than the conventional display (Image b).

The GEE analyses (Table S2a, Table S2b) verified these results with Image (c) consistently having lower odds, for adults (range: 0.13-0.31) and adolescents (range: 0.11-0.32), of being rated favourably compared than Image (b).

### Comparison of ratings between a restricted and conventional display: Image (d) vs. Image (b)

Paired analyses indicated that adults and adolescents consistently rated the fully covered vape and pouch pack display (Image d) less favourably than the conventional display (Image b), see Table S1a and Table S1b.

The GEE analyses (Table S2a, Table S2b) verified these results with Image (d) consistently having lower odds, for adults (range: 0.06-0.19) and adolescents (range: 0.05-0.10), of being rated favourably compared to Image (b).

### Adults’ perceptions on likely product choice from conventional and restricted displays

Around one in ten (11%) adult nicotine users indicated they would not choose any products from Image (c), and 18% would choose none from Image (d) (Figure S1). Compared with Image (b), most remained consistent in their product choice for (c) (83%) and (d) (73%), (Figure S2). For Images (c) and (d) respectively, 4% (n=96) and 6% (n=153) switched to choosing cigarettes/tobacco and, of these, only n=8 and n=20, did not currently smoke. These were adults who currently vaped but had smoked in the past.

### Adolescents who have never vaped: comparison of ratings by susceptibility to vape

Among the 81% (n=1,721, weighted) of adolescents who had never vaped, 40.3% (n=694, weighted) were classed as being susceptible to vape. Logistic regressions, after controlling for demographic and behavioural variables found an association between favourable ratings of conventional displays and likelihood of vaping susceptibility (Table S3). Those giving overall favourable ratings of Images (a) and (b) were more likely to be classed as susceptible: Image (a) AOR=2.73, 95%CI [2.16-3.45] p<0.001; Image (b) AOR=1.89, 95%CI [1.53-2.33] p<0.001.

No statistically significant association was found between susceptibility and favourable ratings of the two restricted displays.

## DISCUSSION

This study examined current and former adult nicotine users’ and adolescents’ responses to conventional and restricted POS displays for vapes and nicotine pouches. The findings show the effect of different displays on perceptions of vapes and vaping. Adults and adolescents rated a large open display, with no age restriction signage or tobacco, more positively across most marketing measures than a conventional display with signage and placed next to tobacco. They consistently rated displays which featured white standardised vape and nicotine pouch packaging or fully covered products more negatively than a conventional display placed next to tobacco. The image of fully covered products, indicative of a display ban, was the most negatively rated overall. When presented with restricted displays, most adults currently using nicotine remained consistent with their product choice. Among adolescents who had never vaped, for the two conventional displays there was an association between favourable ratings and likelihood of vaping susceptibility. No association was found between favourable ratings of the two restricted displays and vaping susceptibility.

Consistent with evidence across consumer goods, the findings highlight how open POS displays of fully-branded vaping and nicotine pouch products positively influence product perceptions and social norms.^7–12^ As a marketing communications tool, the study shows how successful POS displays can be at communicating messages. It demonstrates differences in messaging even between the two conventional displays, with the larger display rated more favourably than the smaller display with age restriction signage and next to tobacco, for all items except ‘makes vaping seem appealing’. One possible explanation with respect to appeal is that adults have been found to view large, brightly coloured displays of vape packaging as targeted towards children and young people.^26^ By focusing specifically on POS displays, the study expands research on retail vape advertising generally,^39^ and vape POS research, which have evidenced associations between display exposure and youth susceptibility to use products.^23–25^ The association between favourable display ratings and adolescent vaping susceptibility found here further suggests that conventional displays could potentially be reinforcing positive vaping attitudes and behaviour.

Less favourable ratings of restricted displays indicate that the removal of POS marketing, either through white standardised packaging or fully covering packs, results in less positive messages being communicated about vaping. Branded packaging is a proven marketing vehicle for attracting attention at POS and delivering product information.^7,9^ Displaying white standardised packs shifted most adult and many adolescent ratings from positive to negative. However, covering packs completely had the greatest shift in ratings. This suggests that banning displays may contribute best to repositioning the current image of vaping and vapes (fun, colourful and youthful),^26,40^ to something more adult-appropriate. It may also be more successful in denormalising vaping and reducing susceptibility to vape (and therefore vaping prevalence) among young people. Notably, for adults’ and adolescents’ assessment of the restricted displays, the highest rated item was ‘makes me think that lots of people vape’, suggesting that the presence of even restricted displays may still give the impression of prevalent vaping, albeit reduced.

The findings have implications for policymakers. A ban on the open display of vapes and other nicotine products may be most effective for reducing visibility and appeal to young people, in line with the UK Government’s objective outlined in their consultation.^2^ There are, however, two important issues to consider. First, is the potential impact on perceptions of vaping harm among adults who may benefit from switching from smoking to vaping, given vaping’s role in smoking cessation. In the current study, display restrictions increased perceptions of harm among adolescents and adults.^41^ Second, a fully covered display had the greatest influence on perceptions of accessibility. Both adults and adolescents rated a fully covered display as ‘makes it difficult for someone like me to buy vapes’. A display ban may discourage impulse purchasing of vapes and nicotine pouches, and purchase attempts among adolescents. In qualitative research with adults who smoked and/or vaped in England and Scotland, some supported a vape display ban to reduce the appeal to young people and felt that that they would overcome any frustration related to not being able to view flavour and device options.^26^ A display ban would have implications for retailers. A study examining the tobacco POS ban in Scotland found that while retailers’ expectations around implementation and impact were negative, retailers reported, post-ban, that implementation was straightforward and concerns about a negative impact on customer transactions were largely unfounded.^42^ There is evidence that retailers are already considering how to make their nicotine product displays compliant with a potential ban, such as concealing existing displays or developing tobacco-style gantries for vapes.^43,44^

### Strengths and limitations

This is the first study to explore responses to different POS displays, using carefully designed, tested, relevant and balanced marketing measures. The study benefits from exploring adolescent and adult current and former nicotine user perspectives, both key groups of interest for policymakers. The study has limitations. Participants rated mock-up images of displays, therefore reactions may differ in real life. The study does not offer insight into how reactions to the restricted displays may change over time. While images included vaping and nicotine pouch products and signage, measures focused on vapes only. Research is needed to explore response to nicotine pouch displays, which are increasing in prominence and size.^28,30^ Product choice questions were hypothetical, and their answers require in-depth qualitative research, however, they provide an indication of reactions to restricting displays from those using tobacco and nicotine products. The surveys used non-probability samples which have issues for generalisability and prevalence estimates.^45^ The samples may not be truly representative, although the adolescent sample was weighted to reflect the UK adolescent population. Recruiting 11 15-year-olds via their parent, online survey administration, and reliance on self-reported data could introduce bias. However, the survey is designed to be completed on any device. Completing on a mobile phone may help protect participants’ privacy. The cross-sectional design does not demonstrate causality in the association between conventional displays and adolescent vaping susceptibility, however, with a large proportion of adolescents susceptible to vaping (42%) this is a concern. Longitudinal research is needed to investigate this association.

### Conclusions

Restricting POS displays could potentially reduce positive messaging around vaping and adolescent vaping susceptibility. Consideration should be given to the potential impact of restrictions on current and former adult nicotine users’ perceptions of harm and accessibility.

## ACKNOWLEDGMENTS

The authors thank YouGov for delivery and management of the survey fieldwork, Jacqui Whyte for facilitating recruitment for the public involvement groups and cognitive interviews, and all the participants who took part in the survey development phase or the survey.

## DATA AVAILABILITY

Participant consent was not obtained for data sharing beyond the scope of the original study.

