## Supplementary material for "Exploring possible point-of-sale restrictions for vapes and nicotine pouches: cross-sectional surveys with adults and adolescents in the UK": Table S1a

### Table S1a. Mean ratings on response to conventional and restricted POS versus 'conventional branded beside covered tobacco': Adults

| | | (a) v (b) Conventional display _ branded vapes & pouches (no tobacco) vs. Conventional display _ branded vapes & pouches beside tobacco (covered) | | | |
|---|---|---|---|---|---|
| | | (a) | (b) | | |
| | | 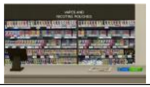 | 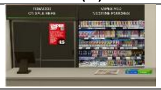 | | |
| | n | Mean [95% CI]<br>SD | Mean [95% CI]<br>SD | Effect Size (r ) # | P Value* |
| **Appraisal** | | | | | |
| Looks unattractive (1)/...attractive (5) | 2851 | 3.66 [3.61, 3.71]<br>*1.34* | 3.41 [3.36, 3.46]<br>*1.27* | 0.30 | <0.001 |
| Does not look eye-catching (1)/...Looks eye-catching (5) | 2851 | 3.78 [3.73, 3.82]<br>*1.28* | 3.50 [3.46, 3.54]<br>*1.19* | 0.30 | <0.001 |
| **Receptivity** | | | | | |
| Makes vaping seem unappealing (1)/...appealing (5) | 2851 | 3.33 [3.29, 3.38]<br>*1.19* | 3.44 [3.40, 3.49]<br>*1.19* | 0.12 | <0.001 |
| Puts me off vaping (1)/...Tempts me to vape (5) | 2851 | 3.38 [3.34, 3.43]<br>*1.22* | 3.24 [3.20, 3.28]<br>*1.16* | 0.22 | <0.001 |
| Makes it feel like vapes are NOT meant for someone like me (1)/... ARE meant (5) | 2851 | 3.30 [3.26, 3.35]<br>*1.18* | 3.18 [3.14, 3.22]<br>*1.12* | 0.18 | <0.001 |
| **Image** | | | | | |
| Makes vaping seem boring (1)/...fun (5) | 2851 | 3.73 [3.69, 3.78]<br>*1.15* | 3.52 [3.48, 3.56]<br>*1.09* | 0.29 | <0.001 |
| Makes me think that vapes are very harmful to health (1)/...not at all harmful (5) | 2851 | 3.65 [3.60, 3.69]<br>*1.18* | 3.46 [3.42, 3.50]<br>*1.14* | 0.25 | <0.001 |
| **Norms** | | | | | |
| Makes me think that it's NOT OK to vape (1)/...it's OK to vape (5) | 2851 | 3.68 [3.64, 3.73]<br>*1.17* | 3.55 [3.50, 3.59]<br>*1.15* | 0.17 | <0.001 |
| Makes me think that hardly anyone vapes (1)/...lots of people vape (5) | 2851 | 4.21 [4.17, 4.25]<br>*1.03* | 3.93 [3.89, 3.97]<br>*1.01* | 0.35 | <0.001 |
| **Functionality** | | | | | |
| Makes it difficult for someone like me to buy vapes (1)/...easy (5) | 2851 | 4.05 [4.01, 4.10]<br>*1.21* | 3.90 [3.86, 3.94]<br>*1.16* | 0.21 | <0.001 |
| **Total score from all 10 items**<br>Minimum = 10, Maximum = 50 | 2851 | 36.8 [36.5, 37.1]<br>*8.81* | 35.1 [34.8, 35.4]<br>*8.47* | 0.40 | <0.001 |

Base: All adults (n=2.851). 'Not sure' responses recoded to midpoint (3). * Wilcoxon signed rank test for significant difference. # Effect Size estimate r, for Wilcoxon signed rank test, r = |Z|/√N. Small effect (r ≥ 0.10); Medium effect (r ≥ 0.30); Large effect (r ≥ 0.50).

Table S1a cont'd. Mean ratings on response to conventional and restricted POS versus 'conventional branded beside covered tobacco': Adults

| | (c) v (b) White vape & pouch packs beside tobacco (covered) vs. Conventional display _ branded vapes & pouches beside tobacco (covered) | | | | | (d) v (b) Fully covered vape & pouch packs beside tobacco (covered) vs. Conventional display _ branded vapes & pouches beside tobacco (covered) | | | | |
|---|---|---|---|---|---|---|---|---|---|---|
| | | (c) | (b) | | | | (d) | (b) | | |
| | | 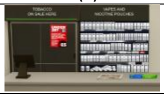 | 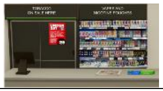 | | | | 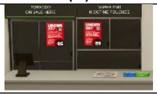 | 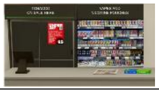 | | |
| | n | Mean [95% CI]<br>SD | Mean [95% CI]<br>SD | Effect Size (r ) # | P Value* | n | Mean [95% CI]<br>SD | Mean [95% CI]<br>SD | Effect Size (r ) # | P Value* |
| **Appraisal** | | | | | | | | | | |
| Looks unattractive (1)/...attractive (5) | 2851 | 2.34 [2.30, 2.38]<br>*1.12* | 3.41 [3.36, 3.46]<br>*1.27* | 0.61 | <0.001 | 2851 | 1.75 [1.71, 1.79]<br>*1.07* | 3.41 [3.36, 3.46]<br>*1.27* | 0.68 | <0.001 |
| Does not look eye-catching (1)/...Looks eye-catching (5) | 2851 | 2.29 [2.25, 2.33]<br>*1.14* | 3.50 [3.46, 3.54]<br>*1.19* | 0.66 | <0.001 | 2851 | 1.61 [1.57, 1.65]<br>*1.04* | 3.50 [3.46, 3.54]<br>*1.19* | 0.73 | <0.001 |
| **Receptivity** | | | | | | | | | | |
| Makes vaping seem unappealing (1)/...appealing (5) | 2851 | 2.39 [2.35, 2.43]<br>*1.11* | 3.44 [3.40, 3.49]<br>*1.19* | 0.61 | <0.001 | 2851 | 1.96 [1.92, 2.00]<br>1.13 | 3.44 [3.40, 3.49]<br>*1.19* | 0.65 | <0.001 |
| Puts me off vaping (1)/...Tempts me to vape (5) | 2851 | 2.59 [2.55, 2.62]<br>*1.03* | 3.24 [3.20, 3.28]<br>*1.16* | 0.52 | <0.001 | 2851 | 2.06 [2.02, 2.10]<br>*1.08* | 3.24 [3.20, 3.28]<br>*1.16* | 0.61 | <0.001 |
| Makes it feel like vapes are NOT meant for someone like me (1)/... ARE meant (5) | 2851 | 2.60 [2.56, 2.64]<br>*1.04* | 3.18 [3.14, 3.22]<br>*1.12* | 0.47 | <0.001 | 2851 | 2.14 [2.1, 2.18]<br>*1.12* | 3.18 [3.14, 3.22]<br>*1.12* | 0.57 | <0.001 |
| **Image** | | | | | | | | | | |
| Makes vaping seem boring (1)/...fun (5) | 2851 | 2.49 [2.45, 2.53]<br>*1.05* | 3.52 [3.48, 3.56]<br>*1.09* | 0.62 | <0.001 | 2851 | 2.02 [1.98, 2.07]<br>*1.13* | 3.52 [3.48, 3.56]<br>*1.09* | 0.66 | <0.001 |
| Makes me think that vapes are very harmful to health (1)/...not at all harmful (5) | 2851 | 2.69 [2.65, 2.73]<br>*1.06* | 3.46 [3.42, 3.50]<br>*1.14* | 0.56 | <0.001 | 2851 | 1.94 [1.90, 1.98]<br>*1.06* | 3.46 [3.42, 3.50]<br>*1.14* | 0.68 | <0.001 |
| **Norms** | | | | | | | | | | |
| Makes me think that it's NOT OK to vape (1)/...it's OK to vape (5) | 2851 | 2.71 [2.67, 2.75]<br>*1.09* | 3.55 [3.50, 3.59]<br>*1.15* | 0.59 | <0.001 | 2851 | 1.87 [1.83, 1.91]<br>*1.08* | 3.55 [3.50, 3.59]<br>*1.15* | 0.70 | <0.001 |
| Makes me think that hardly anyone vapes (1)/...lots of people vape (5) | 2851 | 3.23 [3.19, 3.27]<br>*1.06* | 3.93 [3.89, 3.97]<br>*1.01* | 0.56 | <0.001 | 2851 | 2.54 [2.50, 2.58]<br>*1.18* | 3.93 [3.89, 3.97]<br>*1.01* | 0.67 | <0.001 |
| **Functionality** | | | | | | | | | | |
| Makes it difficult for someone like me to buy vapes (1)/...easy (5) | 2851 | 3.10 [3.06, 3.15]<br>*1.24* | 3.90 [3.86, 3.94]<br>*1.16* | 0.55 | <0.001 | 2851 | 2.28 [2.23, 2.33]<br>*1.31* | 3.9 [3.86, 3.94]<br>*1.16* | 0.68 | <0.001 |
| **Total score from all 10 items**<br>Minimum = 10, Maximum = 50 | 2851 | 26.4 [26.2, 26.7]<br>*7.62* | 35.1 [34.8, 35.4]<br>*8.47* | 0.75 | <0.001 | 2851 | 20.2 [19.9, 20.5]<br>*8.12* | 35.1 [34.8, 35.4]<br>*8.47* | 0.79 | <0.001 |

Base: All adults (n=2.851). 'Not sure' responses recoded to midpoint (3). * Wilcoxon signed rank test for significant difference. # Effect Size estimate r, for Wilcoxon signed rank test, r = |Z|/√N. Small effect (r ≥ 0.10); Medium effect (r ≥ 0.30); Large effect (r ≥ 0.50).
