## Supplementary material for "Exploring possible point-of-sale restrictions for vapes and nicotine pouches: cross-sectional surveys with adults and adolescents in the UK": Table S1b

### Table S1b. Mean ratings on response to conventional and restricted POS versus ‘conventional branded beside covered tobacco’: Adolescents

| | | (a) v (b) Conventional display _ branded vapes & pouches (no tobacco) vs. Conventional display _ branded vapes & pouches beside tobacco (covered) | | | |
|---|---|---|---|---|---|
| | | (a) | (b) | | |
| | | 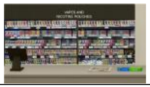 | 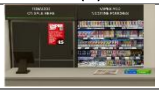 | | |
| | n | Mean [95% CI]<br>SD | Mean [95% CI]<br>SD | Effect Size (r ) # | P Value* |
| **Appraisal** | | | | | |
| Looks unattractive (1)/...attractive (5) | 2123 | 3.58 [3.52, 3.64]<br>*1.47* | 3.16 [3.10, 3.22]<br>*1.40* | 0.45 | <0.001 |
| Does not look eye-catching (1)/...Looks eye-catching (5) | 2123 | 3.61 [3.55, 3.67]<br>*1.43* | 3.22 [3.17, 3.28]<br>*1.33* | 0.39 | <0.001 |
| **Receptivity** | | | | | |
| Makes vaping seem unappealing (1)/...appealing (5) | 2123 | 2.89 [2.83, 2.95]<br>*1.39* | 2.93 [2.87, 2.99]<br>*1.41* | 0.02 | 0.375 |
| Puts me off vaping (1)/...Tempts me to vape (5) | 2123 | 3.12 [3.06, 3.18]<br>*1.42* | 2.80 [2.74, 2.85]<br>*1.34* | 0.38 | <0.001 |
| Makes it feel like vapes are NOT meant for someone like me (1)/... ARE meant (5) | 2123 | 3.15 [3.09, 3.21]<br>*1.42* | 2.84 [2.78, 2.90]<br>*1.36* | 0.36 | <0.001 |
| **Image** | | | | | |
| Makes vaping seem boring (1)/...fun (5) | 2123 | 3.63 [3.57, 3.69]<br>*1.37* | 3.26 [3.21, 3.32]<br>*1.31* | 0.41 | <0.001 |
| Makes me think that vapes are very harmful to health (1)/...not at all harmful (5) | 2123 | 3.56 [3.50, 3.62]<br>*1.41* | 3.18 [3.13, 3.24]<br>*1.36* | 0.40 | <0.001 |
| **Norms** | | | | | |
| Makes me think that it’s NOT OK to vape (1)/...it’s OK to vape (5) | 2123 | 3.43 [3.37, 3.49]<br>*1.41* | 3.12 [3.06, 3.18]<br>*1.39* | 0.34 | <0.001 |
| Makes me think that hardly anyone vapes (1)/...lots of people vape (5) | 2123 | 4.16 [4.11, 4.21]<br>*1.24* | 3.81 [3.76, 3.86]<br>*1.19* | 0.39 | <0.001 |
| **Functionality** | | | | | |
| Makes it difficult for someone like me to buy vapes (1)/...easy (5) | 2123 | 3.54 [3.48, 3.60]<br>*1.41* | 3.15 [3.09, 3.21]<br>*1.37* | 0.39 | <0.001 |
| **Total score from all 10 items**<br>Minimum = 10, Maximum = 50 | 2123 | 34.7 [34.2, 35.1]<br>*10.98* | 31.5 [31.0, 31.9]<br>*10.65* | 0.59 | <0.001 |

Base: All adolescents (n=2.123, weighted). ‘Not sure’ responses recoded to midpoint (3). * Wilcoxon signed rank test for significant difference, conducted on unweighted data. # Effect Size estimate r, for Wilcoxon signed rank test, $r = |Z|/\sqrt{N}$. Small effect ($r \geq 0.10$); Medium effect ($r \geq 0.30$); Large effect ($r \geq 0.50$).

Table S1b cont'd. Mean ratings on response to restricted POS versus 'conventional branded beside covered tobacco': Adolescents

| | (c) v (b) White vape & pouch packs beside tobacco (covered) vs. Conventional display _ branded vapes & pouches beside tobacco (covered) | | | | | (d) v (b) Fully covered vape & pouch packs beside tobacco (covered) vs. Conventional display _ branded vapes & pouches beside tobacco (covered) | | | | |
|---|---|---|---|---|---|---|---|---|---|---|
| | | (c) | (b) | | | | (d) | (b) | | |
| | | 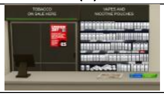 | 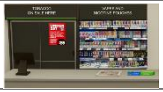 | | | | 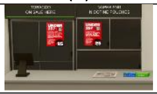 | 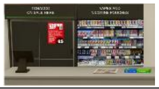 | | |
| | n | Mean [95% CI]<br>SD | Mean [95% CI]<br>SD | Effect Size (r)[#] | P Value* | n | Mean [95% CI]<br>SD | Mean [95% CI]<br>SD | Effect Size (r)[#] | P Value* |
| **Appraisal** | | | | | | | | | | |
| Looks unattractive (1)/...attractive (5) | 2123 | 2.00 [1.95, 2.04]<br>*1.07* | 3.16 [3.10, 3.22]<br>*1.40* | 0.66 | <0.001 | 2123 | 1.41 [1.37, 1.45]<br>*0.87* | 3.16 [3.1, 3.22]<br>*1.40* | 0.71 | <0.001 |
| Does not look eye-catching (1)/...Looks eye-catching (5) | 2123 | 1.98 [1.94, 2.03]<br>*1.10* | 3.22 [3.17, 3.28]<br>*1.33* | 0.68 | <0.001 | 2123 | 1.43 [1.39, 1.46]<br>0.92 | 3.22 [3.17, 3.28]<br>*1.33* | 0.73 | <0.001 |
| **Receptivity** | | | | | | | | | | |
| Makes vaping seem unappealing (1)/...appealing (5) | 2123 | 2.00 [1.95, 2.04]<br>*1.15* | 2.93 [2.87, 2.99]<br>*1.41* | 0.57 | <0.001 | 2123 | 1.56 [1.52, 1.60]<br>*1.00* | 2.93 [2.87, 2.99]<br>*1.41* | 0.64 | <0.001 |
| Puts me off vaping (1)/...Tempts me to vape (5) | 2123 | 2.07 [2.03, 2.12]<br>*1.06* | 2.80 [2.74, 2.85]<br>*1.34* | 0.59 | <0.001 | 2123 | 1.48 [1.44, 1.52]<br>*0.90* | 2.80 [2.74, 2.85]<br>*1.34* | 0.67 | <0.001 |
| Makes it feel like vapes are NOT meant for someone like me (1)/... ARE meant (5) | 2123 | 2.09 [2.04, 2.13]<br>*1.10* | 2.84 [2.78, 2.9]<br>*1.36* | 0.58 | <0.001 | 2123 | 1.43 [1.39, 1.47]<br>*0.88* | 2.84 [2.78, 2.90]<br>*1.36* | 0.68 | <0.001 |
| **Image** | | | | | | | | | | |
| Makes vaping seem boring (1)/...fun (5) | 2123 | 2.13 [2.08, 2.17]<br>*1.06* | 3.26 [3.21, 3.32]<br>*1.31* | 0.67 | <0.001 | 2123 | 1.56 [1.52, 1.60]<br>*0.96* | 3.26 [3.21, 3.32]<br>*1.31* | 0.72 | <0.001 |
| Makes me think that vapes are very harmful to health (1)/...not at all harmful (5) | 2123 | 2.42 [2.37, 2.47]<br>*1.17* | 3.18 [3.13, 3.24]<br>*1.36* | 0.59 | <0.001 | 2123 | 1.55 [1.51, 1.58]<br>*0.93* | 3.18 [3.13, 3.24]<br>*1.36* | 0.71 | <0.001 |
| **Norms** | | | | | | | | | | |
| Makes me think that it's NOT OK to vape (1)/...it's OK to vape (5) | 2123 | 2.34 [2.29, 2.39]<br>*1.20* | 3.12 [3.06, 3.18]<br>*1.39* | 0.58 | <0.001 | 2123 | 1.47 [1.43, 1.51]<br>*0.90* | 3.12 [3.06, 3.18]<br>*1.39* | 0.71 | <0.001 |
| Makes me think that hardly anyone vapes (1)/...lots of people vape (5) | 2123 | 3.04 [2.99, 3.09]<br>*1.20* | 3.81 [3.76, 3.86]<br>*1.19* | 0.60 | <0.001 | 2123 | 2.14 [2.09, 2.19]<br>*1.21* | 3.81 [3.76, 3.86]<br>*1.19* | 0.72 | <0.001 |
| **Functionality** | | | | | | | | | | |
| Makes it difficult for someone like me to buy vapes (1)/...easy (5) | 2123 | 2.44 [2.38, 2.49]<br>*1.23* | 3.15 [3.09, 3.21]<br>*1.37* | 0.56 | <0.001 | 2123 | 1.48 [1.44, 1.52]<br>*0.93* | 3.15 [3.09, 3.21]<br>*1.37* | 0.71 | <0.001 |
| **Total score from all 10 items**<br>Minimum = 10, Maximum = 50 | 2123 | 22.5 [22.2, 22.9]<br>*8.20* | 31.5 [31.0, 31.9]<br>*10.65* | 0.79 | <0.001 | 2123 | 15.5 [15.2, 15.8]<br>*6.88* | 31.5 [31.0, 31.9]<br>*10.65* | 0.82 | <0.001 |

Base: All adolescents (n=2.123, weighted). 'Not sure' responses recoded to midpoint (3). * Wilcoxon signed rank test for significant difference, conducted on unweighted data. [#] Effect Size estimate r, for Wilcoxon signed rank test, r = |Z|/√N. Small effect (r ≥ 0.10); Medium effect (r ≥ 0.30); Large effect (r ≥ 0.50).
