## Supplementary material for "Exploring possible point-of-sale restrictions for vapes and nicotine pouches: cross-sectional surveys with adults and adolescents in the UK": Table S2a

**Table S2a: General estimating equations for binary outcomes: Adults' perceptions of conventional point of sale image (branded vapes and pouches with tobacco covered) v alternative designs: Appraisal**

| | Attractive<br>1= Looks attractive (score 4-5)<br>0= Neutral or unattractive (score ≤3) | | | | Eye-catching<br>1= Looks eye-catching (score 4-5)<br>0= Neutral or not (score ≤3) | | | |
|---|---|---|---|---|---|---|---|---|
| | N | AOR* | 95% CI | P | N | AOR* | 95% CI | P |
| **Style of POS** | | | | | | | | |
| Image (d) Fully covered | 2851 | 0.07 | [0.06, 0.08] | <0.001 | 2851 | 0.06 | [0.05, 0.07] | <0.001 |
| Image (c) White vape & pouch packs (tobacco | 2851 | 0.16 | [0.14, 0.17] | <0.001 | 2851 | 0.13 | [0.11, 0.15] | <0.001 |
| Image (a) Branded vapes & pouches (no tobacco) | 2851 | 1.44 | [1.36, 1.53] | <0.001 | 2851 | 1.49 | [1.40, 1.59] | <0.001 |
| Image (b) Conventional branded display (tobacco covered) | 2851 | Ref | | | 2851 | Ref | | |
| **Gender** | | | | | | | | |
| Female | 5656 | 1.19 | [1.05, 1.35] | 0.01 | 5656 | 1.11 | [0.98, 1.25] | 0.11 |
| Male | 5748 | Ref | | | 5748 | Ref | | |
| **Age group** | | | | | | | | |
| 55+ | 5256 | 0.59 | [0.50, 0.70] | <0.001 | 5256 | 0.46 | [0.39, 0.54] | <0.001 |
| 35-54 | 3588 | 0.67 | [0.56, 0.79] | <0.001 | 3588 | 0.60 | [0.51, 0.72] | <0.001 |
| 18-34 | 2560 | Ref | | | 2560 | Ref | | |
| **Social Grade** | | | | | | | | |
| C2DE | 3488 | 1.02 | [0.89, 1.18] | 0.74 | 3488 | 0.96 | [0.84, 1.11] | 0.60 |
| ABC1 | 7916 | Ref | | | 7916 | Ref | | |
| **Current vaping and/or smoking status** | | | | | | | | |
| Exclusive smoke | 3216 | 0.94 | [0.77, 1.14] | 0.52 | 3216 | 0.97 | [0.80, 1.17] | 0.73 |
| Exclusive vape | 4100 | 1.05 | [0.87, 1.27] | 0.61 | 4100 | 0.98 | [0.81, 1.17] | 0.79 |
| Dual use | 2160 | 1.35 | [1.09, 1.68] | 0.01 | 2160 | 1.28 | [1.03, 1.58] | 0.03 |
| Neither vape nor smoke | 1928 | Ref | | | 1928 | Ref | | |
| **Current pouch use status (used in past 30 days)** | | | | | | | | |
| Current use | 556 | 2.15 | [1.56, 2.97] | <0.001 | 556 | 2.30 | [1.65, 3.23] | <0.001 |
| Does not use | 10848 | Ref | | | 10848 | Ref | | |

* adjusted for all other variables in the model, AOR, adjusted odds ratio; ref, reference category; 95% CI, 95% confidence interval.

**Table S2a Cont'd: General estimating equations for binary outcomes: Adults' perceptions of conventional point of sale image (branded vapes and pouches with tobacco covered) v alternative designs: Receptivity**

| | Appeal<br>1= Makes vaping seem appealing (score 4-5)<br>0= Neutral or unappealing (score ≤3) | | | | Temptation<br>1= Tempts me to vape (score 4-5)<br>0= Neutral or puts off (score ≤3) | | | | Identification<br>1= Makes me think vapes are meant for someone like me (score 4-5)<br>0= Neutral or not (score ≤3) | | | |
|---|---|---|---|---|---|---|---|---|---|---|---|---|
| | **N** | **AOR*** | **95% CI** | **P** | **N** | **AOR*** | **95% CI** | **P** | **N** | **AOR*** | **95% CI** | **P** |
| **Style of POS** | | | | | | | | | | | | |
| Image (d) Fully covered | 2851 | 0.10 | [0.09, 0.12] | <0.001 | 2851 | 0.12 | [0.11, 0.14] | <0.001 | 2851 | 0.19 | [0.17, 0.22] | <0.001 |
| Image (c) White vape & pouch packs (tobacco | 2851 | 0.16 | [0.15, 0.19] | <0.001 | 2851 | 0.27 | [0.24, 0.30] | <0.001 | 2851 | 0.31 | [0.28, 0.34] | <0.001 |
| Image (a) Branded vapes & pouches (no tobacco) | 2851 | 0.79 | [0.74, 0.85] | <0.001 | 2851 | 1.28 | [1.21, 1.35] | <0.001 | 2851 | 1.22 | [1.16, 1.30] | <0.001 |
| Image (b) Conventional branded display (tobacco covered) | 2851 | Ref | | | 2851 | Ref | | | 2851 | Ref | | |
| **Gender** | | | | | | | | | | | | |
| Female | 5656 | 1.00 | [0.89, 1.13] | 0.97 | 5656 | 1.14 | [0.99, 1.32] | 0.06 | 5656 | 1.22 | [1.06, 1.40] | 0.005 |
| Male | 5748 | Ref | | | 5748 | Ref | | | 5748 | Ref | | |
| **Age group** | | | | | | | | | | | | |
| 55+ | 5256 | 0.66 | [0.56, 0.77] | <0.001 | 5256 | 0.47 | [0.39, 0.57] | <0.001 | 5256 | 0.68 | [0.57, 0.82] | <0.001 |
| 35-54 | 3588 | 0.70 | [0.59, 0.82] | <0.001 | 3588 | 0.65 | [0.54, 0.78] | <0.001 | 3588 | 0.73 | [0.61, 0.88] | <0.001 |
| 18-34 | 2560 | Ref | | | 2560 | Ref | | | 2560 | Ref | | |
| **Social Grade** | | | | | | | | | | | | |
| C2DE | 3488 | 0.92 | [0.80, 1.05] | 0.21 | 3488 | 0.97 | [0.82, 1.13] | 0.66 | 3488 | 0.96 | [0.82, 1.12] | 0.60 |
| ABC1 | 7916 | Ref | | | 7916 | Ref | | | 7916 | Ref | | |
| **Current vaping and/or smoking status** | | | | | | | | | | | | |
| Exclusive smoke | 3216 | 0.86 | [0.71, 1.04] | 0.11 | 3216 | 0.89 | [0.71, 1.13] | 0.34 | 3216 | 1.03 | [0.82, 1.28] | 0.83 |
| Exclusive vape | 4100 | 0.84 | [0.70, 1.01] | 0.06 | 4100 | 1.43 | [1.15, 1.77] | 0.001 | 4100 | 1.34 | [1.08, 1.66] | 0.01 |
| Dual use | 2160 | 1.26 | [1.03, 1.54] | 0.02 | 2160 | 1.95 | [1.53, 2.47] | <0.001 | 2160 | 1.77 | [1.40, 2.24] | <0.001 |
| Neither vape nor smoke | 1928 | Ref | | | 1928 | Ref | | | 1928 | Ref | | |
| **Current pouch use status (used in past 30 days)** | | | | | | | | | | | | |
| Current use | 556 | 1.76 | [1.30, 2.39] | <0.001 | 556 | 1.89 | [1.41, 2.55] | <0.001 | 556 | 2.23 | [1.64, 3.02] | <0.001 |
| Does not use | 10848 | Ref | | | 10848 | Ref | | | 10848 | Ref | | |

* adjusted for all other variables in the model, AOR, adjusted odds ratio; ref, reference category; 95% CI, 95% confidence interval.

**Table S2a Cont'd: General estimating equations for binary outcomes: Adults' perceptions of conventional point of sale image (branded vapes and pouches with tobacco covered) v alternative designs: Image**

| | Fun<br>1= Makes vaping seem fun (score 4-5)<br>0= Neutral or boring (score ≤3) | | | | Harm to health<br>1= Makes me think vapes are not at all harmful to health (score 4-5)<br>0= Neutral or harmful (score ≤3) | | | |
|---|---|---|---|---|---|---|---|---|
| | **N** | **AOR*** | **95% CI** | **P** | **N** | **AOR*** | **95% CI** | **P** |
| **Style of POS** | | | | | | | | |
| Image (d) Fully covered | 2851 | 0.10 | [0.08, 0.11] | <0.001 | 2851 | 0.09 | [0.08, 0.11] | <0.001 |
| Image (c) White vape & pouch packs (tobacco | 2851 | 0.16 | [0.14, 0.18] | <0.001 | 2851 | 0.27 | [0.25, 0.30] | <0.001 |
| Image (a) Branded vapes & pouches (no tobacco) | 2851 | 1.38 | [1.30, 1.45] | <0.001 | 2851 | 1.32 | [1.25, 1.39] | <0.001 |
| Image (b) Conventional branded display (tobacco covered) | 2851 | Ref | | | 2851 | Ref | | |
| **Gender** | | | | | | | | |
| Female | 5656 | 1.30 | [1.14, 1.48] | <0.001 | 5656 | 1.19 | [1.04, 1.35] | 0.01 |
| Male | 5748 | Ref | | | 5748 | Ref | | |
| **Age group** | | | | | | | | |
| 55+ | 5256 | 0.55 | [0.47, 0.66] | <0.001 | 5256 | 1.01 | [0.85, 1.20] | 0.90 |
| 35-54 | 3588 | 0.73 | [0.61, 0.86] | <0.001 | 3588 | 1.06 | [0.88, 1.26] | 0.54 |
| 18-34 | 2560 | Ref | | | 2560 | Ref | | |
| **Social Grade** | | | | | | | | |
| C2DE | 3488 | 0.94 | [0.81, 1.08] | 0.37 | 3488 | 0.91 | [0.79, 1.06] | 0.22 |
| ABC1 | 7916 | Ref | | | 7916 | Ref | | |
| **Current vaping and/or smoking status** | | | | | | | | |
| Exclusive smoke | 3216 | 0.80 | [0.65, 0.97] | 0.02 | 3216 | 0.77 | [0.63, 0.94] | 0.01 |
| Exclusive vape | 4100 | 0.73 | [0.60, 0.88] | 0.001 | 4100 | 0.67 | [0.55, 0.81] | <0.001 |
| Dual use | 2160 | 1.09 | [0.87, 1.35] | 0.46 | 2160 | 0.82 | [0.65, 1.02] | 0.07 |
| Neither vape nor smoke | 1928 | Ref | | | 1928 | Ref | | |
| **Current pouch use status (used in past 30 days)** | | | | | | | | |
| Current use | 556 | 2.32 | [1.67, 3.24] | <0.001 | 556 | 1.63 | [1.19, 2.22] | 0.002 |
| Does not use | 10848 | Ref | | | 10848 | Ref | | |

* adjusted for all other variables in the model, AOR, adjusted odds ratio; ref, reference category; 95% CI, 95% confidence interval.

**Table S2a Cont'd: General estimating equations for binary outcomes: Adults' perceptions of conventional point of sale image (branded vapes and pouches with tobacco covered) v alternative designs: Norms**

| | Ok to vape<br>1= Makes me think that it's ok to vape (score 4-5)<br>0= Neutral or does not (score ≤3) | | | | Popularity of vaping<br>1= Makes me think that lots of people vape (score 4-5)<br>0= Neutral or hardly anyone (score ≤3) | | | |
|---|---|---|---|---|---|---|---|---|
| | **N** | **AOR*** | **95% CI** | **P** | **N** | **AOR*** | **95% CI** | **P** |
| **Style of POS** | | | | | | | | |
| Image (d) Fully covered | 2851 | 0.08 | [0.07, 0.09] | <0.001 | 2851 | 0.11 | [0.10, 0.12] | <0.001 |
| Image (c) White vape & pouch packs (tobacco | 2851 | 0.23 | [0.21, 0.25] | <0.001 | 2851 | 0.30 | [0.27, 0.32] | <0.001 |
| Image (a) Branded vapes & pouches (no tobacco) | 2851 | 1.17 | [1.11, 1.24] | <0.001 | 2851 | 1.52 | [1.43, 1.62] | <0.001 |
| Image (b) Conventional branded display (tobacco covered) | 2851 | Ref | | | 2851 | Ref | | |
| **Gender** | | | | | | | | |
| Female | 5656 | 1.21 | [1.07, 1.38] | 0.003 | 5656 | 1.23 | [1.08, 1.39] | 0.001 |
| Male | 5748 | Ref | | | 5748 | Ref | | |
| **Age group** | | | | | | | | |
| 55+ | 5256 | 0.74 | [0.62, 0.87] | <0.001 | 5256 | 0.79 | [0.67, 0.94] | 0.01 |
| 35-54 | 3588 | 0.86 | [0.72, 1.03] | 0.10 | 3588 | 0.86 | [0.72, 1.02] | 0.08 |
| 18-34 | 2560 | Ref | | | 2560 | Ref | | |
| **Social Grade** | | | | | | | | |
| C2DE | 3488 | 0.86 | [0.74, 0.99] | 0.04 | 3488 | 0.97 | [0.85, 1.12] | 0.70 |
| ABC1 | 7916 | Ref | | | 7916 | Ref | | |
| **Current vaping and/or smoking status** | | | | | | | | |
| Exclusive smoke | 3216 | 0.91 | [0.75, 1.12] | 0.38 | 3216 | 0.80 | [0.67, 0.97] | 0.02 |
| Exclusive vape | 4100 | 0.84 | [0.69, 1.03] | 0.09 | 4100 | 0.75 | [0.62, 0.90] | 0.002 |
| Dual use | 2160 | 1.08 | [0.87, 1.35] | 0.47 | 2160 | 0.82 | [0.66, 1.01] | 0.07 |
| Neither vape nor smoke | 1928 | Ref | | | 1928 | Ref | | |
| **Current pouch use status (used in past 30 days)** | | | | | | | | |
| Current use | 556 | 1.85 | [1.35, 2.53] | <0.001 | 556 | 1.70 | [1.25, 2.32] | <0.001 |
| Does not use | 10848 | Ref | | | 10848 | Ref | | |

* adjusted for all other variables in the model, AOR, adjusted odds ratio; ref, reference category; 95% CI, 95% confidence interval.

**Table S2a Cont'd: General estimating equations for binary outcomes: Adults' perceptions of conventional point of sale image (branded vapes and pouches with tobacco covered) v alternative designs: Functionality**

| | Ease of buying<br>1= Makes it easy for someone like me to buy vapes (score 4-5)<br>0= Neutral or difficult (score ≤3) | | | |
|---|---|---|---|---|
| | **N** | **AOR*** | **95% CI** | **P** |
| **Style of POS** | | | | |
| Image (d) Fully covered | 2851 | 0.12 | [0.10, 0.13] | <0.001 |
| Image (c) White vape & pouch packs (tobacco | 2851 | 0.31 | [0.29, 0.34] | <0.001 |
| Image (a) Branded vapes & pouches (no tobacco) | 2851 | 1.24 | [1.18, 1.31] | <0.001 |
| Image (b) Conventional branded display (tobacco covered) | 2851 | Ref | | |
| **Gender** | | | | |
| Female | 5656 | 1.17 | [1.03, 1.33] | 0.02 |
| Male | 5748 | Ref | | |
| **Age group** | | | | |
| 55+ | 5256 | 0.70 | [0.59, 0.82] | <0.001 |
| 35-54 | 3588 | 0.81 | [0.68, 0.95] | 0.01 |
| 18-34 | 2560 | Ref | | |
| **Social Grade** | | | | |
| C2DE | 3488 | 1.01 | [0.88, 1.16] | 0.91 |
| ABC1 | 7916 | Ref | | |
| **Current vaping and/or smoking status** | | | | |
| Exclusive smoke | 3216 | 0.98 | [0.81, 1.19] | 0.85 |
| Exclusive vape | 4100 | 0.98 | [0.82, 1.19] | 0.87 |
| Dual use | 2160 | 1.09 | [0.88, 1.34] | 0.43 |
| Neither vape nor smoke | 1928 | Ref | | |
| **Current pouch use status (used in past 30 days)** | | | | |
| Current use | 556 | 1.73 | [1.26, 2.36] | 0.001 |
| Does not use | 10848 | Ref | | |

* adjusted for all other variables in the model, AOR, adjusted odds ratio; ref, reference category; 95% CI, 95% confidence interval.

**Table S2a: General estimating equations for binary outcomes: Adults' perceptions of conventional point of sale image (branded vapes and pouches with tobacco covered) v alternative designs: Appraisal**

| | Attractive<br>1= Looks attractive (score 4-5)<br>0= Neutral or unattractive (score ≤3) | | | | Eye-catching<br>1= Looks eye-catching (score 4-5)<br>0= Neutral or not (score ≤3) | | | |
|---|---|---|---|---|---|---|---|---|
| | N | AOR* | 95% CI | P | N | AOR* | 95% CI | P |
| **Style of POS** | | | | | | | | |
| Image (d) Fully covered | 2851 | 0.07 | [0.06, 0.08] | <0.001 | 2851 | 0.06 | [0.05, 0.07] | <0.001 |
| Image (c) White vape & pouch packs (tobacco | 2851 | 0.16 | [0.14, 0.17] | <0.001 | 2851 | 0.13 | [0.11, 0.15] | <0.001 |
| Image (a) Branded vapes & pouches (no tobacco) | 2851 | 1.44 | [1.36, 1.53] | <0.001 | 2851 | 1.49 | [1.40, 1.59] | <0.001 |
| Image (b) Conventional branded display (tobacco covered) | 2851 | Ref | | | 2851 | Ref | | |
| **Gender** | | | | | | | | |
| Female | 5656 | 1.19 | [1.05, 1.35] | 0.01 | 5656 | 1.11 | [0.98, 1.25] | 0.11 |
| Male | 5748 | Ref | | | 5748 | Ref | | |
| **Age group** | | | | | | | | |
| 55+ | 5256 | 0.59 | [0.50, 0.70] | <0.001 | 5256 | 0.46 | [0.39, 0.54] | <0.001 |
| 35-54 | 3588 | 0.67 | [0.56, 0.79] | <0.001 | 3588 | 0.60 | [0.51, 0.72] | <0.001 |
| 18-34 | 2560 | Ref | | | 2560 | Ref | | |
| **Social Grade** | | | | | | | | |
| C2DE | 3488 | 1.02 | [0.89, 1.18] | 0.74 | 3488 | 0.96 | [0.84, 1.11] | 0.60 |
| ABC1 | 7916 | Ref | | | 7916 | Ref | | |
| **Current vaping and/or smoking status** | | | | | | | | |
| Exclusive smoke | 3216 | 0.94 | [0.77, 1.14] | 0.52 | 3216 | 0.97 | [0.80, 1.17] | 0.73 |
| Exclusive vape | 4100 | 1.05 | [0.87, 1.27] | 0.61 | 4100 | 0.98 | [0.81, 1.17] | 0.79 |
| Dual use | 2160 | 1.35 | [1.09, 1.68] | 0.01 | 2160 | 1.28 | [1.03, 1.58] | 0.03 |
| Neither vape nor smoke | 1928 | Ref | | | 1928 | Ref | | |
| **Current pouch use status (used in past 30 days)** | | | | | | | | |
| Current use | 556 | 2.15 | [1.56, 2.97] | <0.001 | 556 | 2.30 | [1.65, 3.23] | <0.001 |
| Does not use | 10848 | Ref | | | 10848 | Ref | | |

* adjusted for all other variables in the model, AOR, adjusted odds ratio; ref, reference category; 95% CI, 95% confidence interval.

**Table S2a Cont'd: General estimating equations for binary outcomes: Adults' perceptions of conventional point of sale image (branded vapes and pouches with tobacco covered) v alternative designs: Receptivity**

| | Appeal<br>1= Makes vaping seem appealing (score 4-5)<br>0= Neutral or unappealing (score ≤3) | | | | Temptation<br>1= Tempts me to vape (score 4-5)<br>0= Neutral or puts off (score ≤3) | | | | Identification<br>1= Makes me think vapes are meant for someone like me (score 4-5)<br>0= Neutral or not (score ≤3) | | | |
|---|---|---|---|---|---|---|---|---|---|---|---|---|
| | **N** | **AOR*** | **95% CI** | **P** | **N** | **AOR*** | **95% CI** | **P** | **N** | **AOR*** | **95% CI** | **P** |
| **Style of POS** | | | | | | | | | | | | |
| Image (d) Fully covered | 2851 | 0.10 | [0.09, 0.12] | <0.001 | 2851 | 0.12 | [0.11, 0.14] | <0.001 | 2851 | 0.19 | [0.17, 0.22] | <0.001 |
| Image (c) White vape & pouch packs (tobacco | 2851 | 0.16 | [0.15, 0.19] | <0.001 | 2851 | 0.27 | [0.24, 0.30] | <0.001 | 2851 | 0.31 | [0.28, 0.34] | <0.001 |
| Image (a) Branded vapes & pouches (no tobacco) | 2851 | 0.79 | [0.74, 0.85] | <0.001 | 2851 | 1.28 | [1.21, 1.35] | <0.001 | 2851 | 1.22 | [1.16, 1.30] | <0.001 |
| Image (b) Conventional branded display (tobacco covered) | 2851 | Ref | | | 2851 | Ref | | | 2851 | Ref | | |
| **Gender** | | | | | | | | | | | | |
| Female | 5656 | 1.00 | [0.89, 1.13] | 0.97 | 5656 | 1.14 | [0.99, 1.32] | 0.06 | 5656 | 1.22 | [1.06, 1.40] | 0.005 |
| Male | 5748 | Ref | | | 5748 | Ref | | | 5748 | Ref | | |
| **Age group** | | | | | | | | | | | | |
| 55+ | 5256 | 0.66 | [0.56, 0.77] | <0.001 | 5256 | 0.47 | [0.39, 0.57] | <0.001 | 5256 | 0.68 | [0.57, 0.82] | <0.001 |
| 35-54 | 3588 | 0.70 | [0.59, 0.82] | <0.001 | 3588 | 0.65 | [0.54, 0.78] | <0.001 | 3588 | 0.73 | [0.61, 0.88] | <0.001 |
| 18-34 | 2560 | Ref | | | 2560 | Ref | | | 2560 | Ref | | |
| **Social Grade** | | | | | | | | | | | | |
| C2DE | 3488 | 0.92 | [0.80, 1.05] | 0.21 | 3488 | 0.97 | [0.82, 1.13] | 0.66 | 3488 | 0.96 | [0.82, 1.12] | 0.60 |
| ABC1 | 7916 | Ref | | | 7916 | Ref | | | 7916 | Ref | | |
| **Current vaping and/or smoking status** | | | | | | | | | | | | |
| Exclusive smoke | 3216 | 0.86 | [0.71, 1.04] | 0.11 | 3216 | 0.89 | [0.71, 1.13] | 0.34 | 3216 | 1.03 | [0.82, 1.28] | 0.83 |
| Exclusive vape | 4100 | 0.84 | [0.70, 1.01] | 0.06 | 4100 | 1.43 | [1.15, 1.77] | 0.001 | 4100 | 1.34 | [1.08, 1.66] | 0.01 |
| Dual use | 2160 | 1.26 | [1.03, 1.54] | 0.02 | 2160 | 1.95 | [1.53, 2.47] | <0.001 | 2160 | 1.77 | [1.40, 2.24] | <0.001 |
| Neither vape nor smoke | 1928 | Ref | | | 1928 | Ref | | | 1928 | Ref | | |
| **Current pouch use status (used in past 30 days)** | | | | | | | | | | | | |
| Current use | 556 | 1.76 | [1.30, 2.39] | <0.001 | 556 | 1.89 | [1.41, 2.55] | <0.001 | 556 | 2.23 | [1.64, 3.02] | <0.001 |
| Does not use | 10848 | Ref | | | 10848 | Ref | | | 10848 | Ref | | |

* adjusted for all other variables in the model, AOR, adjusted odds ratio; ref, reference category; 95% CI, 95% confidence interval.

**Table S2a Cont'd: General estimating equations for binary outcomes: Adults' perceptions of conventional point of sale image (branded vapes and pouches with tobacco covered) v alternative designs: Image**

| | Fun<br>1= Makes vaping seem fun (score 4-5)<br>0= Neutral or boring (score ≤3) | | | | Harm to health<br>1= Makes me think vapes are not at all harmful to health (score 4-5)<br>0= Neutral or harmful (score ≤3) | | | |
|---|---|---|---|---|---|---|---|---|
| | **N** | **AOR*** | **95% CI** | **P** | **N** | **AOR*** | **95% CI** | **P** |
| **Style of POS** | | | | | | | | |
| Image (d) Fully covered | 2851 | 0.10 | [0.08, 0.11] | <0.001 | 2851 | 0.09 | [0.08, 0.11] | <0.001 |
| Image (c) White vape & pouch packs (tobacco | 2851 | 0.16 | [0.14, 0.18] | <0.001 | 2851 | 0.27 | [0.25, 0.30] | <0.001 |
| Image (a) Branded vapes & pouches (no tobacco) | 2851 | 1.38 | [1.30, 1.45] | <0.001 | 2851 | 1.32 | [1.25, 1.39] | <0.001 |
| Image (b) Conventional branded display (tobacco covered) | 2851 | Ref | | | 2851 | Ref | | |
| **Gender** | | | | | | | | |
| Female | 5656 | 1.30 | [1.14, 1.48] | <0.001 | 5656 | 1.19 | [1.04, 1.35] | 0.01 |
| Male | 5748 | Ref | | | 5748 | Ref | | |
| **Age group** | | | | | | | | |
| 55+ | 5256 | 0.55 | [0.47, 0.66] | <0.001 | 5256 | 1.01 | [0.85, 1.20] | 0.90 |
| 35-54 | 3588 | 0.73 | [0.61, 0.86] | <0.001 | 3588 | 1.06 | [0.88, 1.26] | 0.54 |
| 18-34 | 2560 | Ref | | | 2560 | Ref | | |
| **Social Grade** | | | | | | | | |
| C2DE | 3488 | 0.94 | [0.81, 1.08] | 0.37 | 3488 | 0.91 | [0.79, 1.06] | 0.22 |
| ABC1 | 7916 | Ref | | | 7916 | Ref | | |
| **Current vaping and/or smoking status** | | | | | | | | |
| Exclusive smoke | 3216 | 0.80 | [0.65, 0.97] | 0.02 | 3216 | 0.77 | [0.63, 0.94] | 0.01 |
| Exclusive vape | 4100 | 0.73 | [0.60, 0.88] | 0.001 | 4100 | 0.67 | [0.55, 0.81] | <0.001 |
| Dual use | 2160 | 1.09 | [0.87, 1.35] | 0.46 | 2160 | 0.82 | [0.65, 1.02] | 0.07 |
| Neither vape nor smoke | 1928 | Ref | | | 1928 | Ref | | |
| **Current pouch use status (used in past 30 days)** | | | | | | | | |
| Current use | 556 | 2.32 | [1.67, 3.24] | <0.001 | 556 | 1.63 | [1.19, 2.22] | 0.002 |
| Does not use | 10848 | Ref | | | 10848 | Ref | | |

* adjusted for all other variables in the model, AOR, adjusted odds ratio; ref, reference category; 95% CI, 95% confidence interval.

**Table S2a Cont'd: General estimating equations for binary outcomes: Adults' perceptions of conventional point of sale image (branded vapes and pouches with tobacco covered) v alternative designs: Norms**

| | Ok to vape<br>1= Makes me think that it's ok to vape (score 4-5)<br>0= Neutral or does not (score ≤3) | | | | Popularity of vaping<br>1= Makes me think that lots of people vape (score 4-5)<br>0= Neutral or hardly anyone (score ≤3) | | | |
|---|---|---|---|---|---|---|---|---|
| | **N** | **AOR*** | **95% CI** | **P** | **N** | **AOR*** | **95% CI** | **P** |
| **Style of POS** | | | | | | | | |
| Image (d) Fully covered | 2851 | 0.08 | [0.07, 0.09] | <0.001 | 2851 | 0.11 | [0.10, 0.12] | <0.001 |
| Image (c) White vape & pouch packs (tobacco | 2851 | 0.23 | [0.21, 0.25] | <0.001 | 2851 | 0.30 | [0.27, 0.32] | <0.001 |
| Image (a) Branded vapes & pouches (no tobacco) | 2851 | 1.17 | [1.11, 1.24] | <0.001 | 2851 | 1.52 | [1.43, 1.62] | <0.001 |
| Image (b) Conventional branded display (tobacco covered) | 2851 | Ref | | | 2851 | Ref | | |
| **Gender** | | | | | | | | |
| Female | 5656 | 1.21 | [1.07, 1.38] | 0.003 | 5656 | 1.23 | [1.08, 1.39] | 0.001 |
| Male | 5748 | Ref | | | 5748 | Ref | | |
| **Age group** | | | | | | | | |
| 55+ | 5256 | 0.74 | [0.62, 0.87] | <0.001 | 5256 | 0.79 | [0.67, 0.94] | 0.01 |
| 35-54 | 3588 | 0.86 | [0.72, 1.03] | 0.10 | 3588 | 0.86 | [0.72, 1.02] | 0.08 |
| 18-34 | 2560 | Ref | | | 2560 | Ref | | |
| **Social Grade** | | | | | | | | |
| C2DE | 3488 | 0.86 | [0.74, 0.99] | 0.04 | 3488 | 0.97 | [0.85, 1.12] | 0.70 |
| ABC1 | 7916 | Ref | | | 7916 | Ref | | |
| **Current vaping and/or smoking status** | | | | | | | | |
| Exclusive smoke | 3216 | 0.91 | [0.75, 1.12] | 0.38 | 3216 | 0.80 | [0.67, 0.97] | 0.02 |
| Exclusive vape | 4100 | 0.84 | [0.69, 1.03] | 0.09 | 4100 | 0.75 | [0.62, 0.90] | 0.002 |
| Dual use | 2160 | 1.08 | [0.87, 1.35] | 0.47 | 2160 | 0.82 | [0.66, 1.01] | 0.07 |
| Neither vape nor smoke | 1928 | Ref | | | 1928 | Ref | | |
| **Current pouch use status (used in past 30 days)** | | | | | | | | |
| Current use | 556 | 1.85 | [1.35, 2.53] | <0.001 | 556 | 1.70 | [1.25, 2.32] | <0.001 |
| Does not use | 10848 | Ref | | | 10848 | Ref | | |

* adjusted for all other variables in the model, AOR, adjusted odds ratio; ref, reference category; 95% CI, 95% confidence interval.

**Table S2a Cont'd: General estimating equations for binary outcomes: Adults' perceptions of conventional point of sale image (branded vapes and pouches with tobacco covered) v alternative designs: Functionality**

| | Ease of buying<br>1= Makes it easy for someone like me to buy vapes (score 4-5)<br>0= Neutral or difficult (score ≤3) | | | |
|---|---|---|---|---|
| | **N** | **AOR*** | **95% CI** | **P** |
| **Style of POS** | | | | |
| Image (d) Fully covered | 2851 | 0.12 | [0.10, 0.13] | <0.001 |
| Image (c) White vape & pouch packs (tobacco | 2851 | 0.31 | [0.29, 0.34] | <0.001 |
| Image (a) Branded vapes & pouches (no tobacco) | 2851 | 1.24 | [1.18, 1.31] | <0.001 |
| Image (b) Conventional branded display (tobacco covered) | 2851 | Ref | | |
| **Gender** | | | | |
| Female | 5656 | 1.17 | [1.03, 1.33] | 0.02 |
| Male | 5748 | Ref | | |
| **Age group** | | | | |
| 55+ | 5256 | 0.70 | [0.59, 0.82] | <0.001 |
| 35-54 | 3588 | 0.81 | [0.68, 0.95] | 0.01 |
| 18-34 | 2560 | Ref | | |
| **Social Grade** | | | | |
| C2DE | 3488 | 1.01 | [0.88, 1.16] | 0.91 |
| ABC1 | 7916 | Ref | | |
| **Current vaping and/or smoking status** | | | | |
| Exclusive smoke | 3216 | 0.98 | [0.81, 1.19] | 0.85 |
| Exclusive vape | 4100 | 0.98 | [0.82, 1.19] | 0.87 |
| Dual use | 2160 | 1.09 | [0.88, 1.34] | 0.43 |
| Neither vape nor smoke | 1928 | Ref | | |
| **Current pouch use status (used in past 30 days)** | | | | |
| Current use | 556 | 1.73 | [1.26, 2.36] | 0.001 |
| Does not use | 10848 | Ref | | |

* adjusted for all other variables in the model, AOR, adjusted odds ratio; ref, reference category; 95% CI, 95% confidence interval.
