## Supplementary material for "Exploring possible point-of-sale restrictions for vapes and nicotine pouches: cross-sectional surveys with adults and adolescents in the UK": Table S2b

**Table S2b: General estimating equations for binary outcomes: Adolescents' perceptions of conventional point of sale image (branded vapes and pouches with tobacco covered) v alternative designs: Appraisal**

| | Attractive<br>1= Looks attractive (score 4-5)<br>0= Neutral or unattractive (score ≤3) | | | | Eye-catching<br>1= Looks eye-catching (score 4-5)<br>0= Neutral or not (score ≤3) | | | |
|---|---|---|---|---|---|---|---|---|
| | N | AOR* | 95% CI | P | N | AOR* | 95% CI | P |
| **Style of POS** | | | | | | | | |
| Image (d) Fully covered | 2102 | 0.05 | [0.04, 0.06] | <0.001 | 2102 | 0.06 | [0.05, 0.07] | <0.001 |
| Image (c) White vape & pouch packs (tobacco | 2102 | 0.11 | [0.10, 0.13] | <0.001 | 2102 | 0.13 | [0.11, 0.15] | <0.001 |
| Image (a) Branded vapes & pouches (no tobacco) | 2102 | 1.85 | [1.72, 1.99] | <0.001 | 2102 | 1.88 | [1.74, 2.04] | <0.001 |
| Image (b) Conventional branded display (tobacco covered) | 2102 | Ref | | | 2102 | Ref | | |
| **Gender** | | | | | | | | |
| Female | 3872 | 1.06 | [0.91, 1.24] | 0.46 | 3872 | 0.97 | [0.84, 1.13] | 0.72 |
| Male | 4536 | Ref | | | 4536 | Ref | | |
| **Age group** | | | | | | | | |
| 17 | 1224 | 1.13 | [0.87, 1.45] | 0.36 | 1224 | 0.92 | [0.72, 1.16] | 0.47 |
| 15-16 | 2444 | 1.11 | [0.90, 1.37] | 0.32 | 2444 | 1.01 | [0.83, 1.24] | 0.91 |
| 13-14 | 2416 | 1.39 | [1.13, 1.72] | 0.002 | 2416 | 1.16 | [0.95, 1.42] | 0.14 |
| 11-12 | 2324 | Ref | | | 2324 | Ref | | |
| **Social Grade** | | | | | | | | |
| C2DE | 2244 | 1.07 | [0.90, 1.27] | 0.47 | 2244 | 1.04 | [0.88, 1.23] | 0.67 |
| ABC1 | 6164 | Ref | | | 6164 | Ref | | |
| **Current vaping and/or smoking status** | | | | | | | | |
| Exclusive smoke | 48 | 0.98 | [0.38, 2.49] | 0.96 | 48 | 0.96 | [0.35, 2.63] | 0.94 |
| Exclusive vape | 480 | 2.55 | [1.82, 3.56] | <0.001 | 480 | 2.37 | [1.70, 3.29] | <0.001 |
| Dual use | 92 | 4.77 | [2.26, 10.1] | <0.001 | 92 | 4.32 | [2.00, 9.34] | <0.001 |
| Neither vape nor smoke | 7788 | Ref | | | 7788 | Ref | | |
| **Current pouch use status (used in past 30 days)** | | | | | | | | |
| Current use | 124 | 3.60 | [1.56, 8.30] | 0.003 | 124 | 4.96 | [2.20, 11.17] | <0.001 |
| Does not use | 8284 | Ref | | | 8284 | Ref | | |

* adjusted for all other variables in the model, AOR, adjusted odds ratio; ref, reference category; 95% CI, 95% confidence interval.

Valid cases = 8408; Missing cases=84, excluded due to missing data on one or more variables.

**Table S2b Cont'd: General estimating equations for binary outcomes: Adolescents' perceptions of conventional point of sale image (branded vapes and pouches with tobacco covered) v alternative designs: Receptivity**

| | Appeal 1= Makes vaping seem appealing (score 4-5) 0= Neutral or unappealing (score ≤3) | | | | Temptation 1= Tempts me to vape (score 4-5) 0= Neutral or puts off (score ≤3) | | | | Identification 1= Makes me think vapes are meant for someone like me (score 4-5) 0= Neutral or not (score ≤3) | | | |
|---|---|---|---|---|---|---|---|---|---|---|---|---|
| | N | AOR* | 95% CI | P | N | AOR* | 95% CI | P | N | AOR* | 95% CI | P |
| **Style of POS** | | | | | | | | | | | | |
| Image (d) Fully covered | 2102 | 0.10 | [0.08, 0.12] | <0.001 | 2102 | 0.09 | [0.07, 0.11] | <0.001 | 2102 | 0.07 | [0.06, 0.09] | <0.001 |
| Image (c) White vape & pouch packs (tobacco | 2102 | 0.21 | [0.18, 0.24] | <0.001 | 2102 | 0.23 | [0.20, 0.27] | <0.001 | 2102 | 0.23 | [0.20, 0.26] | <0.001 |
| Image (a) Branded vapes & pouches (no tobacco) | 2102 | 0.90 | [0.82, 0.98] | 0.01 | 2102 | 1.69 | [1.57, 1.81] | <0.001 | 2102 | 1.63 | [1.52, 1.76] | <0.001 |
| Image (b) Conventional branded display (tobacco covered) | 2102 | Ref | | | 2102 | Ref | | | 2102 | Ref | | |
| **Gender** | | | | | | | | | | | | |
| Female | 3872 | 1.05 | [0.90, 1.23] | 0.51 | 3872 | 1.07 | [0.90, 1.27] | 0.42 | 3872 | 1.00 | [0.85, 1.19] | 0.98 |
| Male | 4536 | Ref | | | 4536 | Ref | | | 4536 | Ref | | |
| **Age group** | | | | | | | | | | | | |
| 17 | 1224 | 0.94 | [0.73, 1.21] | 0.64 | 1224 | 1.12 | [0.86, 1.47] | 0.41 | 1224 | 1.23 | [0.93, 1.61] | 0.14 |
| 15-16 | 2444 | 1.15 | [0.94, 1.41] | 0.19 | 2444 | 1.07 | [0.85, 1.34] | 0.58 | 2444 | 1.34 | [1.06, 1.68] | 0.01 |
| 13-14 | 2416 | 1.13 | [0.92, 1.39] | 0.24 | 2416 | 1.24 | [0.99, 1.56] | 0.06 | 2416 | 1.42 | [1.13, 1.78] | 0.003 |
| 11-12 | 2324 | Ref | | | 2324 | Ref | | | 2324 | Ref | | |
| **Social Grade** | | | | | | | | | | | | |
| C2DE | 2244 | 1.18 | [0.99, 1.40] | 0.06 | 2244 | 1.18 | [0.97, 1.43] | 0.09 | 2244 | 1.03 | [0.85, 1.24] | 0.79 |
| ABC1 | 6164 | Ref | | | 6164 | Ref | | | 6164 | Ref | | |
| **Current vaping and/or smoking status** | | | | | | | | | | | | |
| Exclusive smoke | 48 | 0.97 | [0.32, 3.00] | 0.96 | 48 | 0.32 | [0.05, 1.99] | 0.22 | 48 | 0.67 | [0.22, 2.05] | 0.49 |
| Exclusive vape | 480 | 2.33 | [1.74, 3.11] | <0.001 | 480 | 4.39 | [3.17, 6.07] | <0.001 | 480 | 3.46 | [2.46, 4.85] | <0.001 |
| Dual use | 92 | 5.57 | [2.65, 11.70] | <0.001 | 92 | 6.53 | [3.23, 13.18] | <0.001 | 92 | 4.39 | [1.95, 9.88] | <0.001 |
| Neither vape nor smoke | 7788 | Ref | | | 7788 | Ref | | | 7788 | Ref | | |
| **Current pouch use status (used in past 30 days)** | | | | | | | | | | | | |
| Current use | 124 | 3.66 | [1.80, 7.43] | <0.001 | 124 | 1.41 | [0.70, 2.82] | 0.33 | 124 | 2.14 | [1.03, 4.48] | 0.04 |
| Does not use | 8284 | Ref | | | 8284 | Ref | | | 8284 | Ref | | |

* adjusted for all other variables in the model, AOR, adjusted odds ratio; ref, reference category; 95% CI, 95% confidence interval.

Valid cases = 8408; Missing cases=84, excluded due to missing data on one or more variables.

**Table S2b Cont'd: General estimating equations for binary outcomes: Adolescents' perceptions of conventional point of sale image (branded vapes and pouches with tobacco covered) v alternative designs: Image**

| | Fun<br>1= Makes vaping seem fun (score 4-5)<br>0= Neutral or boring (score ≤3) | | | | Harm to health<br>1= Makes me think vapes are not at all harmful to health (score 4-5)<br>0= Neutral or harmful (score ≤3) | | | |
|---|---|---|---|---|---|---|---|---|
| | **N** | **AOR*** | **95% CI** | **P** | **N** | **AOR*** | **95% CI** | **P** |
| **Style of POS** | | | | | | | | |
| Image (d) Fully covered | 2102 | 0.05 | [0.04, 0.07] | <0.001 | 2102 | 0.05 | [0.04, 0.07] | <0.001 |
| Image (c) White vape & pouch packs (tobacco | 2102 | 0.12 | [0.10, 0.13] | <0.001 | 2102 | 0.26 | [0.24, 0.30] | <0.001 |
| Image (a) Branded vapes & pouches (no tobacco) | 2102 | 1.75 | [1.63, 1.88] | <0.001 | 2102 | 1.68 | [1.56, 1.80] | <0.001 |
| Image (b) Conventional branded display (tobacco covered) | 2102 | Ref | | | 2102 | Ref | | |
| **Gender** | | | | | | | | |
| Female | 3872 | 0.91 | [0.78, 1.07] | 0.25 | 3872 | 0.98 | [0.84, 1.15] | 0.85 |
| Male | 4536 | Ref | | | 4536 | Ref | | |
| **Age group** | | | | | | | | |
| 17 | 1224 | 1.01 | [0.79, 1.31] | 0.91 | 1224 | 1.02 | [0.80, 1.32] | 0.86 |
| 15-16 | 2444 | 1.14 | [0.93, 1.40] | 0.22 | 2444 | 1.15 | [0.94, 1.42] | 0.18 |
| 13-14 | 2416 | 1.19 | [0.97, 1.47] | 0.10 | 2416 | 1.09 | [0.88, 1.34] | 0.43 |
| 11-12 | 2324 | Ref | | | 2324 | Ref | | |
| **Social Grade** | | | | | | | | |
| C2DE | 2244 | 1.03 | [0.86, 1.23] | 0.74 | 2244 | 0.80 | [0.67, 0.95] | 0.01 |
| ABC1 | 6164 | Ref | | | 6164 | Ref | | |
| **Current vaping and/or smoking status** | | | | | | | | |
| Exclusive smoke | 48 | 0.71 | [0.33, 1.53] | 0.38 | 48 | 0.19 | [0.03, 1.39] | 0.10 |
| Exclusive vape | 480 | 1.91 | [1.39, 2.63] | <0.001 | 480 | 1.82 | [1.33, 2.49] | <0.001 |
| Dual use | 92 | 2.49 | [1.10, 5.61] | 0.03 | 92 | 1.12 | [0.55, 2.29] | 0.75 |
| Neither vape nor smoke | 7788 | Ref | | | 7788 | Ref | | |
| **Current pouch use status (used in past 30 days)** | | | | | | | | |
| Current use | 124 | 5.14 | [2.31, 11.44] | <0.001 | 124 | 0.53 | [0.29, 0.99] | 0.05 |
| Does not use | 8284 | Ref | | | 8284 | Ref | | |

* adjusted for all other variables in the model, AOR, adjusted odds ratio; ref, reference category; 95% CI, 95% confidence interval.

Valid cases = 8408; Missing cases=84, excluded due to missing data on one or more variables.

**Table S2b Cont'd: General estimating equations for binary outcomes: Adolescents' perceptions of conventional point of sale image (branded vapes and pouches with tobacco covered) v alternative designs: Norms**

| | Ok to vape<br>1= Makes me think that it's ok to vape (score 4-5)<br>0= Neutral or does not (score ≤3) | | | | Popularity of vaping<br>1= Makes me think that lots of people vape (score 4-5)<br>0= Neutral or hardly anyone (score ≤3) | | | |
|---|---|---|---|---|---|---|---|---|
| | **N** | **AOR*** | **95% CI** | **P** | **N** | **AOR*** | **95% CI** | **P** |
| **Style of POS** | | | | | | | | |
| Image (d) Fully covered | 2102 | 0.06 | [0.05, 0.07] | <0.001 | 2102 | 0.08 | [0.07, 0.09] | <0.001 |
| Image (c) White vape & pouch packs (tobacco | 2102 | 0.27 | [0.24, 0.30] | <0.001 | 2102 | 0.27 | [0.25, 0.30] | <0.001 |
| Image (a) Branded vapes & pouches (no tobacco) | 2102 | 1.53 | [1.42, 1.64] | <0.001 | 2102 | 1.69 | [1.57, 1.83] | <0.001 |
| Image (b) Conventional branded display (tobacco covered) | 2102 | Ref | | | 2102 | Ref | | |
| **Gender** | | | | | | | | |
| Female | 3872 | 1.00 | [0.85, 1.17] | 0.98 | 3872 | 1.02 | [0.88, 1.19] | 0.75 |
| Male | 4536 | Ref | | | 4536 | Ref | | |
| **Age group** | | | | | | | | |
| 17 | 1224 | 1.09 | [0.84, 1.40] | 0.53 | 1224 | 1.09 | [0.86, 1.39] | 0.46 |
| 15-16 | 2444 | 1.11 | [0.90, 1.37] | 0.33 | 2444 | 1.19 | [0.98, 1.45] | 0.08 |
| 13-14 | 2416 | 1.19 | [0.96, 1.47] | 0.11 | 2416 | 1.19 | [0.98, 1.45] | 0.08 |
| 11-12 | 2324 | Ref | | | 2324 | Ref | | |
| **Social Grade** | | | | | | | | |
| C2DE | 2244 | 0.92 | [0.77, 1.09] | 0.33 | 2244 | 0.92 | [0.78, 1.09] | 0.32 |
| ABC1 | 6164 | Ref | | | 6164 | Ref | | |
| **Current vaping and/or smoking status** | | | | | | | | |
| Exclusive smoke | 48 | 0.42 | [0.10, 1.87] | 0.26 | 48 | 0.27 | [0.09, 0.79] | 0.02 |
| Exclusive vape | 480 | 2.49 | [1.82, 3.41] | <0.001 | 480 | 1.48 | [1.09, 2.02] | 0.01 |
| Dual use | 92 | 5.66 | [2.40, 13.3] | <0.001 | 92 | 1.74 | [0.82, 3.66] | 0.15 |
| Neither vape nor smoke | 7788 | Ref | | | 7788 | Ref | | |
| **Current pouch use status (used in past 30 days)** | | | | | | | | |
| Current use | 124 | 2.01 | [1.01, 3.98] | 0.05 | 124 | 1.44 | [0.72, 2.90] | 0.30 |
| Does not use | 8284 | Ref | | | 8284 | Ref | | |

* adjusted for all other variables in the model, AOR, adjusted odds ratio; ref, reference category; 95% CI, 95% confidence interval.

Valid cases = 8408; Missing cases=84, excluded due to missing data on one or more variables.

**Table S2b Cont'd: General estimating equations for binary outcomes: Adolescents' perceptions of conventional point of sale image (branded vapes and pouches with tobacco covered) v alternative designs: Functionality**

| | Ease of buying<br>1= Makes it easy for someone like me to buy vapes (score 4-5)<br>0= Neutral or difficult (score ≤3) | | | |
|---|---|---|---|---|
| | **N** | **AOR*** | **95% CI** | **P** |
| **Style of POS** | | | | |
| Image (d) Fully covered | 2102 | 0.06 | [0.05, 0.07] | <0.001 |
| Image (c) White vape & pouch packs (tobacco | 2102 | 0.32 | [0.29, 0.35] | <0.001 |
| Image (a) Branded vapes & pouches (no tobacco) | 2102 | 1.77 | [1.64, 1.90] | <0.001 |
| Image (b) Conventional branded display (tobacco covered) | 2102 | Ref | | |
| **Gender** | | | | |
| Female | 3872 | 1.01 | [0.86, 1.18] | 0.92 |
| Male | 4536 | Ref | | |
| **Age group** | | | | |
| 17 | 1224 | 1.66 | [1.29, 2.13] | <0.001 |
| 15-16 | 2444 | 1.48 | [1.19, 1.83] | <0.001 |
| 13-14 | 2416 | 1.40 | [1.13, 1.74] | 0.002 |
| 11-12 | 2324 | Ref | | |
| **Social Grade** | | | | |
| C2DE | 2244 | 0.98 | [0.81, 1.17] | 0.79 |
| ABC1 | 6164 | Ref | | |
| **Current vaping and/or smoking status** | | | | |
| Exclusive smoke | 48 | 0.19 | [0.02, 1.42] | 0.10 |
| Exclusive vape | 480 | 2.37 | [1.70, 3.32] | <0.001 |
| Dual use | 92 | 2.37 | [1.06, 5.30] | 0.04 |
| Neither vape nor smoke | 7788 | Ref | | |
| **Current pouch use status (used in past 30 days)** | | | | |
| Current use | 124 | 2.47 | [1.15, 5.30] | 0.02 |
| Does not use | 8284 | Ref | | |

* adjusted for all other variables in the model, AOR, adjusted odds ratio; ref, reference category; 95% CI, 95% confidence interval.
Valid cases = 8408; Missing cases=84, excluded due to missing data on one or more variables.
