## Supplementary figures and images for "Exploring possible point-of-sale restrictions for vapes and nicotine pouches: cross-sectional surveys with adults and adolescents in the UK"

### Figure S1

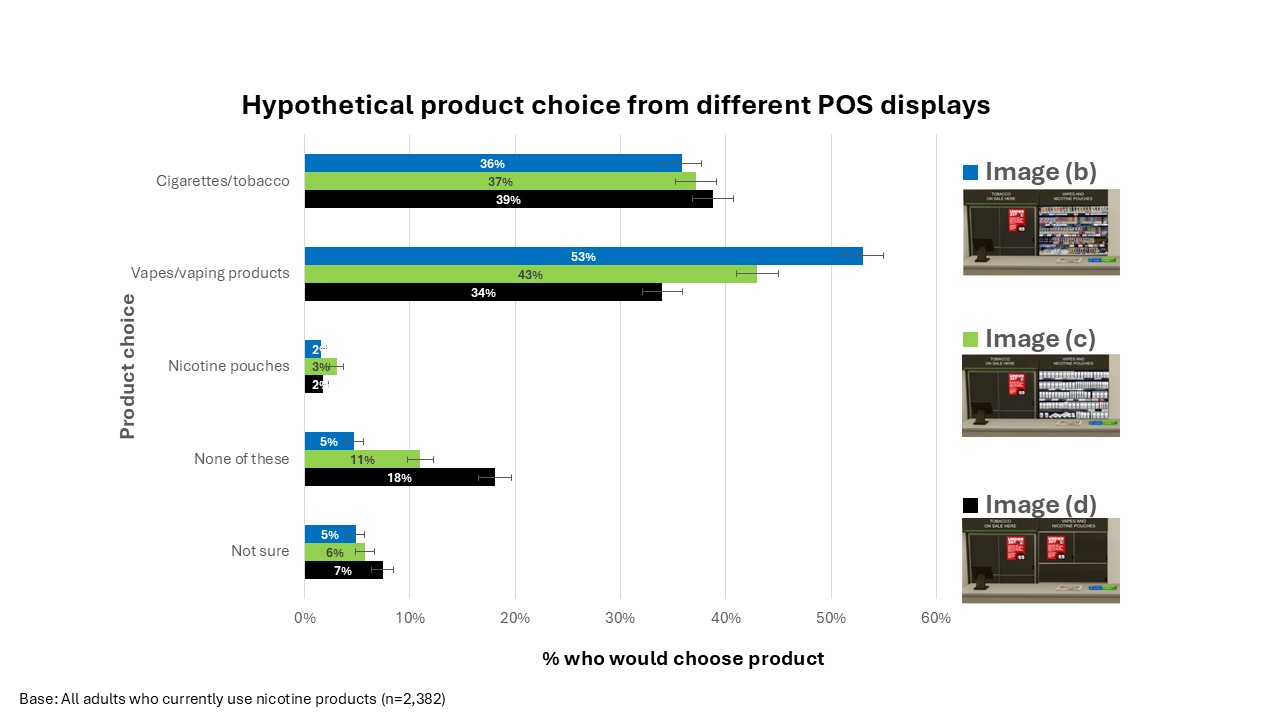

### Figure S2

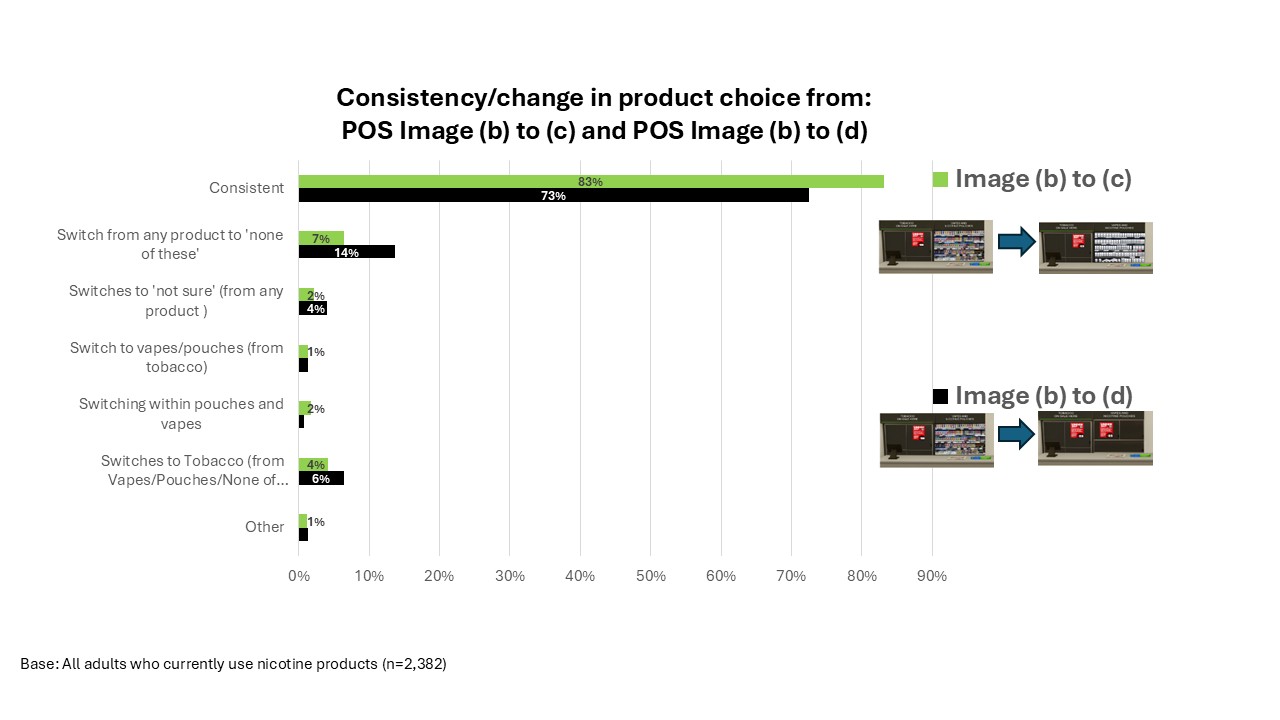
