## Supplementary material for "Exploring possible point-of-sale restrictions for vapes and nicotine pouches: cross-sectional surveys with adults and adolescents in the UK": Table S3

### Table S3: Logistic regression: Association between overall rating of display images and adolescents' susceptibility to vape

| | (a) Conventional display _ branded vapes & pouches (no tobacco) 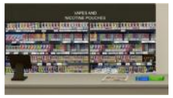 **1= Susceptible (n=685)** [#] **0= Not susceptible (n=956)** | | | | (b) Conventional display _ branded vapes & pouches beside tobacco (covered) 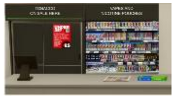 **1= Susceptible (n=685)** [#] **0= Not susceptible (n=956)** | | | |
|---|---|---|---|---|---|---|---|---|
| | **N** | **AOR*** | **N** | **AOR*** | **N** | **AOR*** | **95% CI** | **P** |
| **Peer vaping** | | | | <0.001 | | | | <0.001 |
| Any friends vape at least weekly | 702 | 2.88 | [2.26, 3.68] | <0.001 | 702 | 3.06 | [2.40, 3.88] | <0.001 |
| Not sure | 123 | 1.43 | [0.94, 2.17] | 0.091 | 123 | 1.43 | [0.95, 2.16] | 0.086 |
| No friends vape at least weekly | 816 | Ref | | | 816 | Ref | | |
| **Sibling vaping** | | | | 0.029 | | | | 0.056 |
| Any siblings vape at all nowadays | 130 | 1.63 | [1.10, 2.41] | 0.014 | 130 | 1.55 | [1.05, 2.28] | 0.027 |
| Not sure | 40 | 0.72 | [0.36, 1.45] | 0.361 | 40 | 0.75 | [0.38, 1.50] | 0.416 |
| No siblings who vape | 1471 | Ref | | | 1471 | Ref | | |
| **Parental vaping** | | | | 0.005 | | | | 0.001 |
| Either or both parent(s) vape | 255 | 1.55 | [1.16, 2.08] | 0.003 | 255 | 1.63 | [1.23, 2.18] | <0.001 |
| Not sure | 10 | 2.87 | [0.67, 12.32] | 0.156 | 10 | 3.29 | [0.79, 13.7] | 0.102 |
| Neither parent vapes | 1376 | Ref | | | 1376 | Ref | | |
| **Gender** | | | | | | | | |
| Female | 767 | 0.82 | [0.66, 1.01] | 0.066 | 767 | 0.82 | [0.67, 1.01] | 0.066 |
| Male | 874 | Ref | | | 874 | Ref | | |
| **Age group** | | | | <0.001 | | | | <0.001 |
| 17 | 199 | 0.85 | [0.65, 1.12] | 0.246 | 199 | 0.83 | [0.63, 1.08] | 0.163 |
| 15-16 | 435 | 0.53 | [0.41, 0.69] | <0.001 | 435 | 0.53 | [0.41, 0.69] | <0.001 |
| 13-14 | 504 | 0.48 | [0.34, 0.68] | <0.001 | 504 | 0.49 | [0.35, 0.69] | <0.001 |
| 11-12 | 503 | Ref | | | 503 | Ref | | |
| **Social Grade** | | | | | | | | |
| C2DE | 409 | 0.84 | [0.65, 1.08] | 0.166 | 409 | 0.83 | [0.65, 1.06] | 0.136 |
| ABC1 | 1232 | Ref | | | 1232 | Ref | | |
| **Overall rating of display image (across 10 items)** | | | | | | | | |
| Overall favourable rating of display image | 1056 | 2.73 | [2.16, 3.45] | <0.001 | 830 | 1.89 | [1.53, 2.33] | <0.001 |
| Neutral or unfavourable overall rating | 585 | Ref | | | 811 | Ref | | |
| Model χ2 | | | 231.614 | | | | 191.964 | |
| df, p value | | | 12 | <0.001 | | | 12 | <0.001 |
| Nagelkerke $R^2$ | | | 0.177 | | | | 0.149 | |

* adjusted for all other variables in the model. AOR, adjusted odds ratio; ref, reference category; 95% CI, 95% confidence interval. [#] unweighted

Valid cases =1641, missing cases excluded due to missing data on one or more variables=82. Base: young people who have never vaped.

Bonferroni adjustment accounting for repeated logistic regressions results in critical α= 0.0125.

**Table S3 Cont'd : Logistic regression: Association between overall rating of display images and adolescents' susceptibility to vape**

| | (c) White vape & pouch packs beside tobacco (covered) | | | | (d) Fully covered vape & pouch packs beside tobacco (covered) | | | |
|---|---|---|---|---|---|---|---|---|
| | 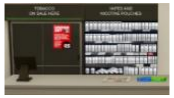 | | | | 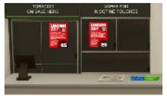 | | | |
| | **1= Susceptible (n=685) #** | | | | **1= Susceptible (n=685) #** | | | |
| | **0= Not susceptible (n=956)** | | | | **0= Not susceptible (n=956)** | | | |
| | **N** | **AOR*** | **N** | **AOR*** | **N** | **AOR*** | **95% CI** | **P** |
| **Peer vaping** | | | | <0.001 | | | | <0.001 |
| Any friends vape at least weekly | 702 | 3.22 | [2.54, 4.08] | <0.001 | 702 | 3.31 | [2.61, 4.20] | <0.001 |
| Not sure | 123 | 1.48 | [0.98, 2.22] | 0.060 | 123 | 1.48 | [0.99, 2.22] | 0.058 |
| No friends vape at least weekly | 816 | Ref | | | 816 | Ref | | |
| **Sibling vaping** | | | | 0.061 | | | | 0.059 |
| Any siblings vape at all nowadays | 130 | 1.55 | [1.05, 2.28] | 0.026 | 130 | 1.55 | [1.06, 2.28] | 0.025 |
| Not sure | 40 | 0.79 | [0.40, 1.56] | 0.496 | 40 | 0.79 | [0.40, 1.57] | 0.502 |
| No siblings who vape | 1471 | Ref | | | 1471 | Ref | | |
| **Parental vaping** | | | | <0.001 | | | | <0.001 |
| Either or both parent(s) vape | 255 | 1.67 | [1.25, 2.22] | <0.001 | 255 | 1.66 | [1.25, 2.21] | <0.001 |
| Not sure | 10 | 3.30 | [0.80, 13.72] | 0.100 | 10 | 3.37 | [0.80, 14.14] | 0.097 |
| Neither parent vapes | 1376 | Ref | | | 1376 | Ref | | |
| **Gender** | | | | | | | | |
| Female | 767 | 0.82 | [0.67, 1.01] | 0.061 | 767 | 0.81 | [0.66, 1.00] | 0.053 |
| Male | 874 | Ref | | | 874 | Ref | | |
| **Age group** | | | | <0.001 | | | | <0.001 |
| 17 | 199 | 0.84 | [0.64, 1.09] | 0.191 | 199 | 0.85 | [0.66, 1.11] | 0.245 |
| 15-16 | 435 | 0.53 | [0.41, 0.68] | <0.001 | 435 | 0.52 | [0.40, 0.67] | <0.001 |
| 13-14 | 504 | 0.48 | [0.34, 0.68] | <0.001 | 504 | 0.48 | [0.34, 0.67] | <0.001 |
| <0.00111-12 | 503 | Ref | | | 503 | Ref | | |
| **Social Grade** | | | | | | | | |
| C2DE | 409 | 0.82 | [0.65, 1.05] | 0.119 | 409 | 0.81 | [0.64, 1.04] | 0.094 |
| ABC1 | 1232 | Ref | | | 1232 | Ref | | |
| **Overall rating of display image (across 10 items)** | | | | | | | | |
| Overall favourable rating of display image | 223 | 1.46 | [1.08, 1.97] | 0.013 | 41 | 0.92 | [0.48, 1.77] | 0.807 |
| Neutral or unfavourable overall rating | 1418 | Ref | | | 1600 | Ref | | |
| Model χ2 | | | 162.422 | | | | 156.33 | |
| df, p value | | | 12 | <0.001 | | | 12 | <0.001 |
| Nagelkerke $R^2$ | | | 0.127 | | | | 0.122 | |

* adjusted for all other variables in the model. AOR, adjusted odds ratio; ref, reference category; 95% CI, 95% confidence interval. # unweighted

Valid cases =1641, missing cases excluded due to missing data on one or more variables=82. Base: young people who have never vaped.

Bonferroni adjustment accounting for repeated logistic regressions results in critical α= 0.0125.
